# Comparative magnitude and persistence of SARS-CoV-2 vaccination responses on a population level in Germany

**DOI:** 10.1101/2021.12.01.21266960

**Authors:** Alex Dulovic, Barbora Kessel, Manuela Harries, Matthias Becker, Julia Ortmann, Johanna Griesbaum, Jennifer Jüngling, Daniel Junker, Pilar Hernandez, Daniela Gornyk, Stephan Glöckner, Vanessa Melhorn, Stefanie Castell, Jana-Kristin Heise, Yvonne Kemmling, Torsten Tonn, Kerstin Frank, Thomas Illig, Norman Klopp, Neha Warikoo, Angelika Rath, Christina Suckel, Anne Ulrike Marzian, Nicole Grupe, Philipp D. Kaiser, Bjoern Traenkle, Ulrich Rothbauer, Tobias Kerrinnes, Gérard Krause, Berit Lange, Nicole Schneiderhan-Marra, Monika Strengert

## Abstract

**Background:** While SARS-CoV-2 vaccinations were successful in decreasing COVID-19 caseloads, recent increases in SARS-CoV-2 infections have led to questions about duration and quality of the subsequent immune response. While numerous studies have been published on immune responses triggered by vaccination, these often focused on the initial peak response generated in specific population subgroups (e.g. healthcare workers or immunocompromised individuals) and have often only examined the effects of one or two different immunisation schemes.

**Methods and Findings:** We analysed serum samples from participants of a large German seroprevalence study (MuSPAD) who had received all available vaccines and dose schedules (mRNA-1273, BNT162b2, AZD1222, Ad26.CoV2S.2 or a combination of AZD1222 plus either mRNA-1273 or BNT162b2). Antibody titers against various SARS-CoV-2 antigens and ACE2 binding inhibition against SARS-CoV-2 wild-type and the Alpha, Beta, Gamma and Delta variants of concern were analysed using a previously published multiplex immunoassay MULTICOV-AB and an ACE2-RBD competition assay. Among the different vaccines and their dosing regimens, homologous mRNA-based or heterologous prime-boost vaccination produced significantly higher antibody responses than vector-based homologous vaccination. Ad26.CoV2S.2 performance was significantly reduced, even compared to AZD1222, with 91.67% of samples being considered non-responsive forACE2 binding inhibition. mRNA-based vaccination induced a higher ratio of RBD- and S1-targeting antibodies than vector-based vaccination, which resulted in an increased proportion of S2-targeting antibodies. Previously infected individuals had a robust immune response once vaccinated, regardless of which vaccine they received. When examining antibody kinetics post-vaccination after homologous immunisation regimens, both titers and ACE2 binding inhibition peaked approximately 28 days post-vaccination and then decreased as time increased.

**Conclusions:** As one of the first and largest population-based studies to examine vaccine responses for all currently available immunisation schemes in Germany, we found that homologous mRNA or heterologous vaccination elicited the highest immune responses. The high percentage of non-responders for Ad26.CoV2.S requires further investigation and suggests that a booster dose with an mRNA-based vaccine may be necessary. The high responses seen in recovered and vaccinated individuals could aid future dose allocation, should shortages arise for certain manufacturers. Given the role of RBD- and S1-specific antibodies in neutralising SARS-CoV-2, their relative over-representation after mRNA vaccination may explain why mRNA vaccines have an increased efficacy compared to vector-based formulations. Further investigation on these differences will be of particular interest for vaccine development and efficacy, especially for the next-generation of vector-based vaccines.

## 1. Introduction

In response to the global SARS-CoV-2 pandemic, multiple vaccines have been developed, tested and licensed for use within record time (1-4). As vaccination coverage became more widespread at the beginning of 2021, countries experienced a reduction in SARS-CoV-2 infections (5, 6), although case numbers have again begun to increase in recent months due to spread among and by unvaccinated individuals (7) as well as longevity-related reductions in vaccine protection (8-11). Although a measurable correlate of protection that either prevents SARS-CoV-2 infection or limits COVID-19 disease progression is not yet defined, sufficient levels of neutralizing antibodies are assumed to be a key element (12, 13). As in most other countries, the German national vaccination strategy (until June 7^th^ 2021) was based on prioritization by occupation, underlying medical conditions or advanced age. Currently, 56.8 million German residents are reported to be completely vaccinated (68.3% coverage), with a further 2.4 million having so far received one dose. The majority of doses administered based on delivery numbers in Germany are BNT162b2 from Pfizer (77.0%), followed by Astra Zeneca AZD1222 (11.3%), Moderna’s mRNA-1273 (8.7%) and Janssen’s single-shot Ad26.CoV2.S (3.0%; impfdashboard.de and rki.de as of November 25^th^ 2021). However, based on a lack of efficacy data from phase III clinical trials, the German Standing Committee on Vaccination (STIKO) recommended AZD1222 only for use in those below the age of 60. Following reports of moderate to severe thrombocytopenia and atypical thrombosis cases after AZD1222 vaccination in spring 2021 (14-16), temporary suspensions and eligibility restrictions were not only enacted in Germany (on March 15^th^ 2021) but in 12 other EU member states (17). Administration of AZD1222 was resumed by the 1^st^ of April 2021 in Germany, however only for those above the age of 60 or after an individual risk analysis. Individuals who had received a first dose of AZD1222 and were below the age of 60 were instead offered a mRNA-based vaccine as second dose which resulted in a heterologous prime-boost vaccination scheme (18). Although these “mix and match” approaches were not covered by the initial licensing terms, it has by now been shown that they result in a more robust humoral and cell-mediated immune response compared to the homologous AZD1222 immunisation (19, 20). While multiple studies have so far investigated vaccine-induced responses, predominantly in at-risk groups such as dialysis or transplant recipients (21, 22), groups with increased exposure risk such as health care workers (23-25) or as part of the initial clinical efficacy trials which in general enrol healthier than average populations (26), we report immunological vaccination response data from the general adult population. By using samples from a population-based seroprevalence study (MuSPAD), which assessed SARS-CoV-2 seroprevalence from July 2020 to August 2021 in eight regions in Germany (27), we examined the dynamics of vaccine-induced humoral responses using MULTICOV-AB (28) and an ACE2-RBD competition assay (29) to analyse ACE2 binding inhibition.

## 2. Methods

### 2.1 MuSPAD study recruitment

Vaccination responses were analysed in participants of the <u>Mu</u>lti-local and <u>s</u>erial cross-sectional <u>p</u>revalence study on <u>a</u>ntibodies against SARS-CoV-2 in <u>G</u>ermany (MuSPAD) study, a nationwide population-based SARS-CoV-2 seroprevalence study (27) from July 2020 to August 2021. The study was approved by the Ethics Committee of the Hannover Medical School (9086_BO_S_2020). MuSPAD participants were recruited by age- and gender-stratified random sampling based on records from the respective local residents’ registration offices. Study locations in eight regions across Germany were selected in spring 2020 based on differing epidemic activity at that time. In addition to the successive cross-sectional study design, certain study locations were sampled longitudinally within a 3-4 month interval. At the study centre, following written informed consent, all eligible participants (>18 years) were subject to a standardised computer-based interview using the digital health tool PIA (Prospective Monitoring and Management-App) to gather basic sociodemographic data, information on pre-existing medical conditions including a previously confirmed SARS-CoV-2 infection or a SARS-CoV-2 vaccination, once it became available in Germany in late December 2020. Information about SARS-CoV-2 infections or vaccinations are self-reported. After serum was obtained by venipuncture from a serum gel S-Monovette (Sarstedt), samples were aliquoted in Matrix 2D Barcoded Screw Top Tubes (Thermo Scientific) at the Institute of Transfusion Medicine and Immunohematology and frozen at −20°C before being transported on dry ice to the Hannover Unified Biobank (Germany). After registration and quality control, one serum aliquot was shipped to the Natural and Medical Sciences Institute (Reutlingen, Germany) where they were stored at −80°C until analysis.

### 2.2 Study design and eligibility

Our study contains a total of 1821 samples from 1731 MuSPAD participants which were divided into three subgroups to examine different aspects of the vaccine-induced humoral response. Based on our inclusion criteria, individual samples can be part of several subgroups.

1. Individuals who received a homologous or heterologous full two-dose vaccination with AZD1222, BNT162b2 and mRNA-1273 or the one-dose vaccine Ad26.CoV2.S with a blood sample taken at least 7 days but no more than 65 days post the last vaccination (hereon referred to as “mix and match sample cohort”)
2. Individuals who donated one blood sample following a two-dose homologous vaccination with BNT162b2 or mRNA-1273 within the defined time frames of day 5 to 12, day 26 to 30, day 54 to 58, day 94 to 103, day 129 to 146 or day 176 to 203 after the second dose to monitor antibody kinetics (hereon referred to as “time point sample cohort”)
3. Individuals with paired blood samples taken at two separate successive time points where the first sample had to be taken a minimum of seven days after the second homologous dose of BNT162b2 (hereon referred to as “longitudinal sample cohort”).

All samples originated from the following locations where the MUSPAD study had previously been scheduled to take place and were collected from January-August 2021: Aachen (Städteregion), Magdeburg (Stadtkreis), Osnabrück (Stadt-und Landkreis), Chemnitz (Stadtkreis) or Landkreis Vorpommern-Greifswald. A flow chart to illustrate sample selection form the entire MuSPAD cohort can be found in **Fig. S1**. Basic sociodemographic information and details of comorbidities (hypertension, cardiovascular disease, diabetes, lung disease, immunosuppression, cancer) for each group are provided in more detail in **Table 1** and **Table S1**. Apart from the homologous BNT162b2 samples which are part of our mix and match sample cohort, the maximum available sample number meeting the specified criteria in groups 1-3 was used. For the homologous BNT162b2 vaccination samples within our mix and match sample cohort, we applied a random selection from the entire available sample pool of BNT162b2 vaccinees who took part in the MuSPAD study to select 771 sera. Individuals with a previous SARS-CoV-2 infection either defined by a positive SARS-CoV-2 PCR or antigen test result, or a nucleocapsid IgG normalisation ratio above 1 are listed separately (hereon referred to as “recovered”) within the mix and match sample cohort. Additional sample eligibility criteria were having a complete vaccination record (manufacturer and vaccination dates) and information on age and gender as part of the participant’s metadata.

**Table 1.**
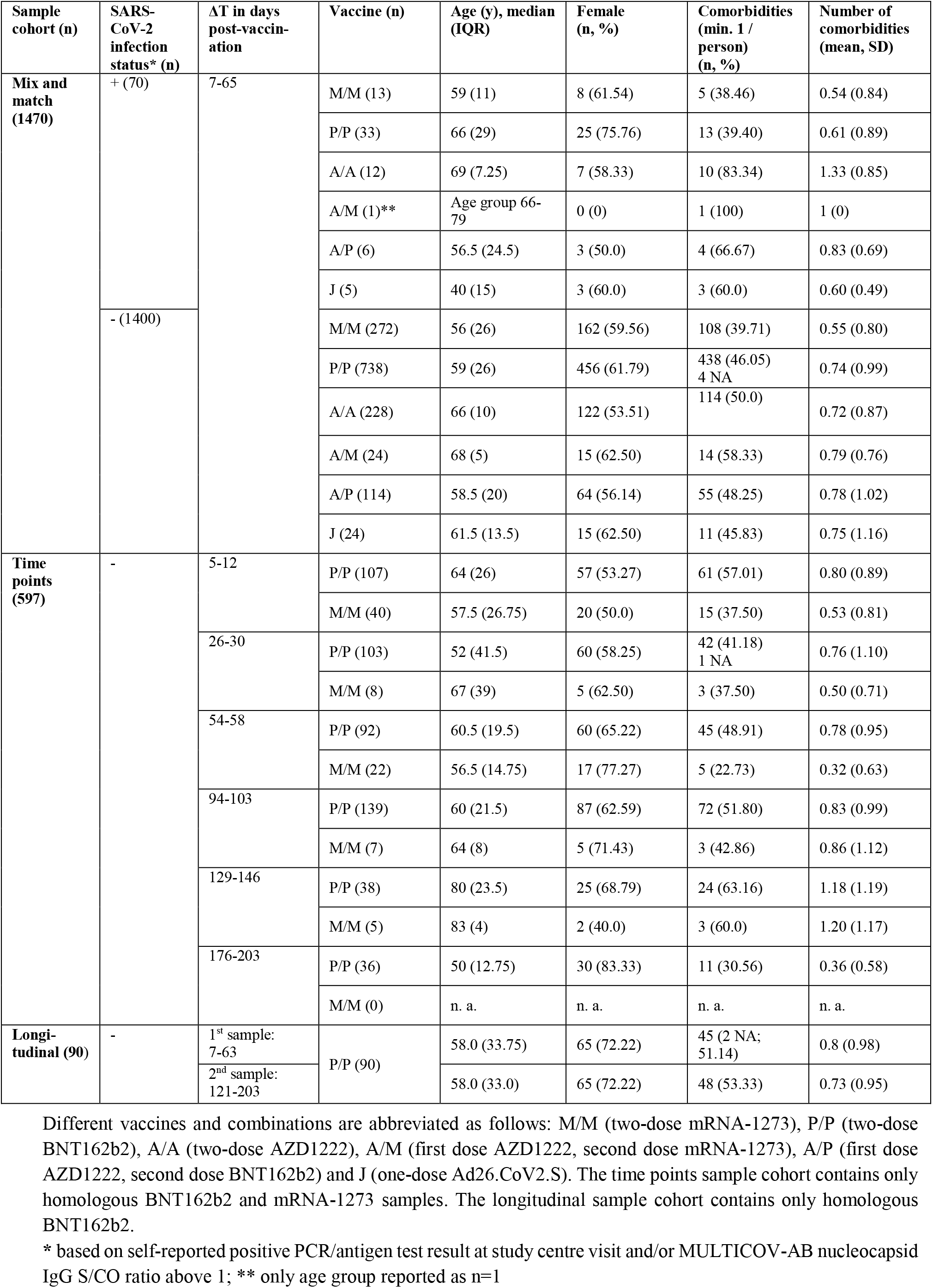
Demographics of study population (n. a.: not applicable; NA: not available).

### 2.3 MULTICOV-AB

Vaccine-induced humoral responses were analysed using MULTICOV-AB (28), a previously published multiplex immunoassay that includes both antigens of SARS-CoV-2 (e.g. Spike, Receptor Binding Domain (RBD), S1 domain, S2 domain and nucleocapsid) and the endemic coronaviruses (OC43, HKU1, NL63 and 229E). Samples were processed using an automated platform on a Beckman Coulter i7 pipetting robot as previously described (30). Briefly, samples were thawed at room temperature, vortexed and then centrifuged at 2000 g for 3 mins to pellet any cell debris within the sample. Samples were then opened using a LabElite DeCapper SL (Hamilton Company). Opened sample matrix racks were then loaded into the pipetting robot, where the sample was diluted 1:200 in assay buffer, before being combined in a 384-well plate and mixed 1:1 with 1x bead mix (see **Table S2** for antigen panel), resulting in a final dilution of 1:400. Samples were then incubated in a Thermomixer (Eppendorf) for 2 h at 1400 rpm, 20°C, in darkness. Following this initial incubation, samples were washed to remove unbound antibodies using an automated magnetic plate washer (Biotek). Bound IgG was detected by adding R-phycoerythrin labelled goat-anti-human IgG (3 µg/mL; #109-116-098, Jackson Immunoresearch Labs) and incubating for a further 45 mins at 1400 rpm, 20°C, in darkness. Following a further washing step, beads were resuspended in 100 µl of wash buffer, shaken for 1 min at 1400 rpm and then measured once on a FLEXMAP 3D instrument (Luminex Corporation) using the following settings: Timeout 100 sec, Gate 7500-15000, Reporter Gain: Standard PMT, 40 events. 3 quality control (QC) samples were included in octuplicate per plate. Any sample that failed QC was remeasured. Raw median fluorescence intensity (MFI) values were normalised to a QC sample for all antigens as in (24, 31).

### 2.4 ACE2-RBD competition assay

To enable high-throughput screening of ACE2-RBD binding inhibition in the presence of sera, a previously established ACE2-RBD competition assay (29) was automated on a Beckmann Coulter i7 pipetting robot. 1:20 previously diluted samples from MULTICOV-AB were diluted 1:200 in ACE2 buffer (29) containing 150 ng/mL biotinylated ACE2. Samples were then mixed 1:1 with 1x VoC (Variant of Concern) bead mix containing RBDs of SARS-CoV-2 wild-type and the Alpha, Beta, Gamma, Delta VoCs (**Table S3**), resulting in a final dilution of 1:400. Samples were then incubated in a Thermomixer for 2 hours at 1400 rpm, 20°C, in darkness. Following this initial incubation, samples were washed to remove unbound ACE2 using an automated magnetic plate washer. ACE2 was detected using R-phycoerythrin labelled streptavidin (2 µg/mL, #SAPE-001, Moss) by incubating the bead-sample mix for a further 45 mins at 1400 rpm, 20°C, in darkness. Following a further washing step, beads were resuspended in 100 µl of wash buffer, shaken for 1 min at 1400 rpm and then measured once on a FLEXMAP 3D instrument using the following settings: Timeout 100 sec, Gate 7500-15000, Reporter Gain: Standard PMT, 40 events. As controls, 12 blank wells, 10 wells with 150 ng/mL ACE2 alone and 10 wells with an ACE2 QC sample were included. ACE2 binding inhibition was calculated as percentage ACE2 inhibition as in (29) with 100% indicating maximum ACE2 binding inhibition and 0% no ACE2 binding inhibition. Samples with an ACE2 binding inhibition less than 20% are classified as non-responders (29).

### 2.5 Data analysis and statistics

Initial results collation and matching to metadata was done in Excel 2016 and R 4.1.0 (32).

For pair-wise comparisons of titres and ACE2 binding inhibition between vaccination schemes within our mix and max sample cohort, we used a two-sided Brunner-Munzel/generalised Wilcoxon test (33) with a significance level of 0.05 as part of the lawstat package (34). In each comparison of two vaccination schemes, the test assesses if a titre (or ACE2 binding inhibition) tends to larger (smaller) values under one vaccination scheme in comparison to the other. Where indicated, we adjusted for multiple testing by using Holm’s procedure (33) to control the family-wise error rate to be below 0.05. To investigate the impact of age, sex, comorbidities and time post-vaccination on the ACE2 binding inhibition between the different vaccination schemes, we used a normal linear mixed model for logit-transformed ACE2 binding inhibition. Negative measurement values were replaced by 0.001 to enable the transformation. The model included additive effects of age, sex, time post-vaccination (peak response period: 7-27 days vs plateau response period: 28-65 days) and comorbidities (cardiovascular disease, hypertension, diabetes, lung disease and cancer/immunosuppression (which were combined to a binary indicator based on low sample numbers). The model further included a random effect defined by the variable “plate number” to account for dependencies due to the measurement procedure, and allowed for heteroscedastic variances for younger (<=70) and older (>70) ages and vaccination types. REML estimation was implemented using the lme function (nlme library (35)). Statistical testing was based on the asymptotic normality of the estimates. As part of a sensitivity analysis, we extended the model with interaction terms between each confounder and the time post-vaccination, allowing for possibly differing effects in the peak (7-27 days) and plateau (28-65 days) period after the last vaccination. Since the effects of the considered covariates were allowed to differ between the vaccination schemes, we analysed only four vaccination schemes (BNT162b2/BNT162b2, mRNA-1273/mRNA-1273, AZD1222/BNT162b2, AZD1222/AZD1222) with a sufficient sample size in the mix and match study cohort. Four individuals with BNT162b2/BNT162b2 vaccination with missing comorbidity metadata were excluded from this analysis. The described statistical comparison of vaccination schemes within the mix and max cohort was performed after the exclusion of recovered individuals. To assess the impact of a previous SARS-CoV-2 infection on RBD antibody titres and wild-type ACE2 binding inhibition among the different vaccination schemes, we also used a two-sided Brunner-Munzel test.

To generate a heat map for comparing antigen-specific antibodies formation across different vaccination schemes within the mix and max sample cohort, normalised antibody responses were initially scaled using the function “z-score”, before being plotted as a heat map. To evaluate longitudinal changes in antibody response and ACE2 binding inhibition within our longitudinal sample cohort, changes from T1 to T2 were calculated using log2 fold change. Any increase in titre or binding is represented by a positive value, while decreases in titre or binding are represented by negative values.

Data visualisation was done in RStudio (Version 1.2.5001 running R version 3.6.1). Additional packages “gplots” (34) and “beeswarm” (35) were used for specific displays. Graphs were exported from RStudio and further edited in Inkscape (Version 0.92.4) to generate final figures.

## 3. Results

First, we examined differences in humoral responses between individuals who received homologous or heterologous immunisation schemes within our mix and match sample cohort where vaccine dose distribution is similar to the German vaccine coverage. Using MULTICOV-AB, we compared vaccination-induced antibody titres generated against the full-length Spike trimer, RBD, S1 and S2 domains and found that mRNA-based homologous vaccinations induced a greater Spike (median normalised MFI mRNA-1273 13.78, BNT162b2 12.49, AZD1222 5.68, Ad26.CoV2.S 3.65) RBD (median normalised MFI mRNA-1273 29.12, BNT162b2 24.89, AZD1222 9.61, Ad26.CoV2.S 5.25) and S1 response (median normalised MFI mRNA-1273 195.9, BNT162b2 139.8, AZD1222 56.40, Ad26.CoV2.S 10.14) than vector-based ones **(Fig. 1)**. When comparing between the two vector-based vaccinations, the two-dose immunisation with AZD1222 resulted in higher titres than the one-dose Ad26.CoV2.S from Janssen. For mRNA vaccines, Moderna’s mRNA-1273 produced a significantly higher response than Pfizer’s BNT162b2 (p-values <0.001, **Table S5**). Heterologous dose vaccination schemes resulted in comparable titres (for Spike and RBD) as homologous mRNA-vaccinated regimens among our study group independent of the origin of the second dose (Spike normalised MFI AZD/mRNA-1273 13.59, AZD/BNT162b2 13.27, RBD normalised MFI AZD/mRNA-1273 28.17, AZD/BNT162b2 25.93). Heterologous titres were in addition significantly higher than those after a homologous AZD1222 two-dose immunisation (p-values <0.001, **Table S5**). In line with their lower titres, serological non-responder rate (defined as a Signal to Cutoff ratio (S/CO) below 1 for either Spike or RBD) was highest for vector-based homologous vaccination schemes (**Table 2**).

**Table 2.**
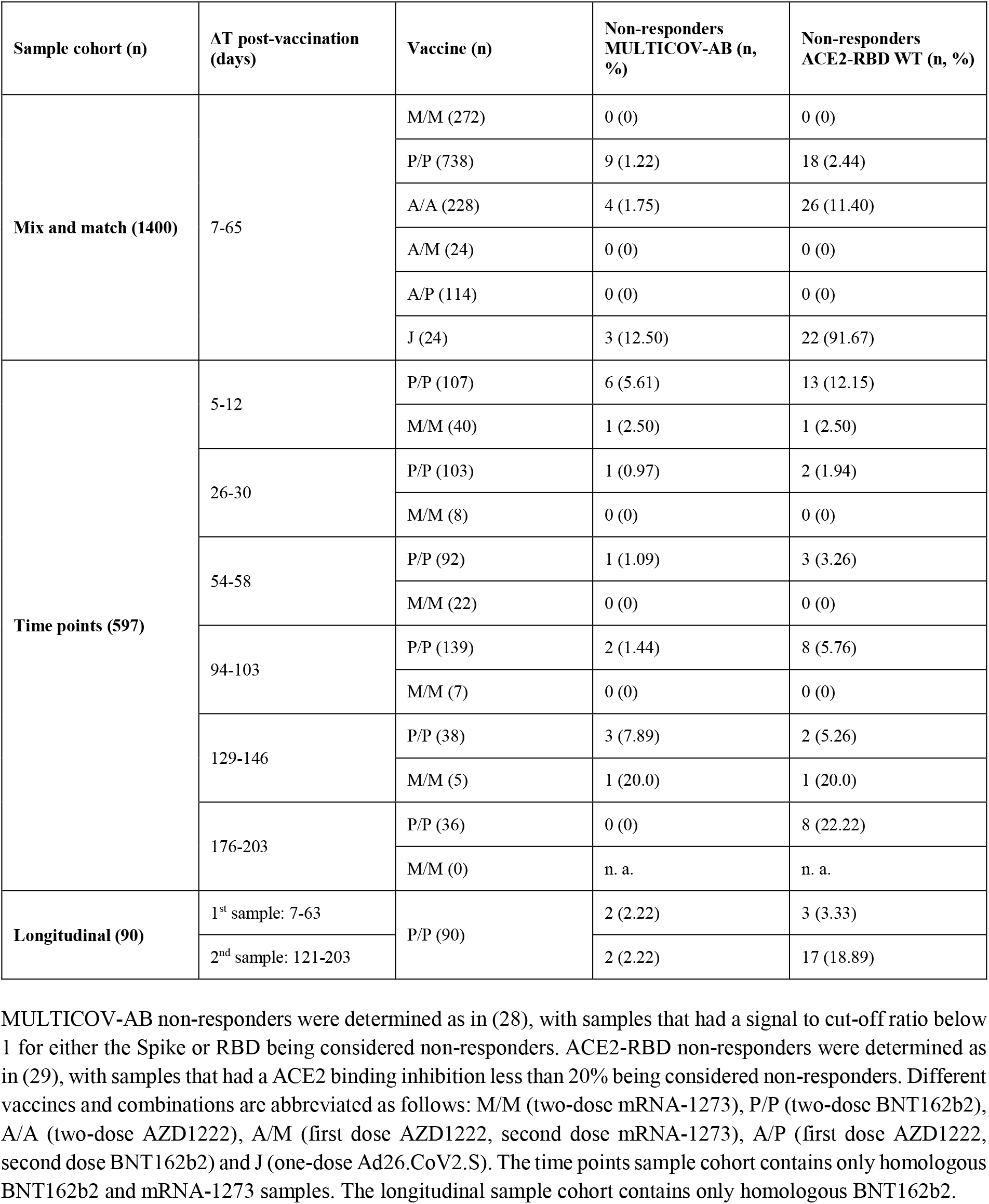
Vaccine non-responder rates across study population.

**Fig. 1.**
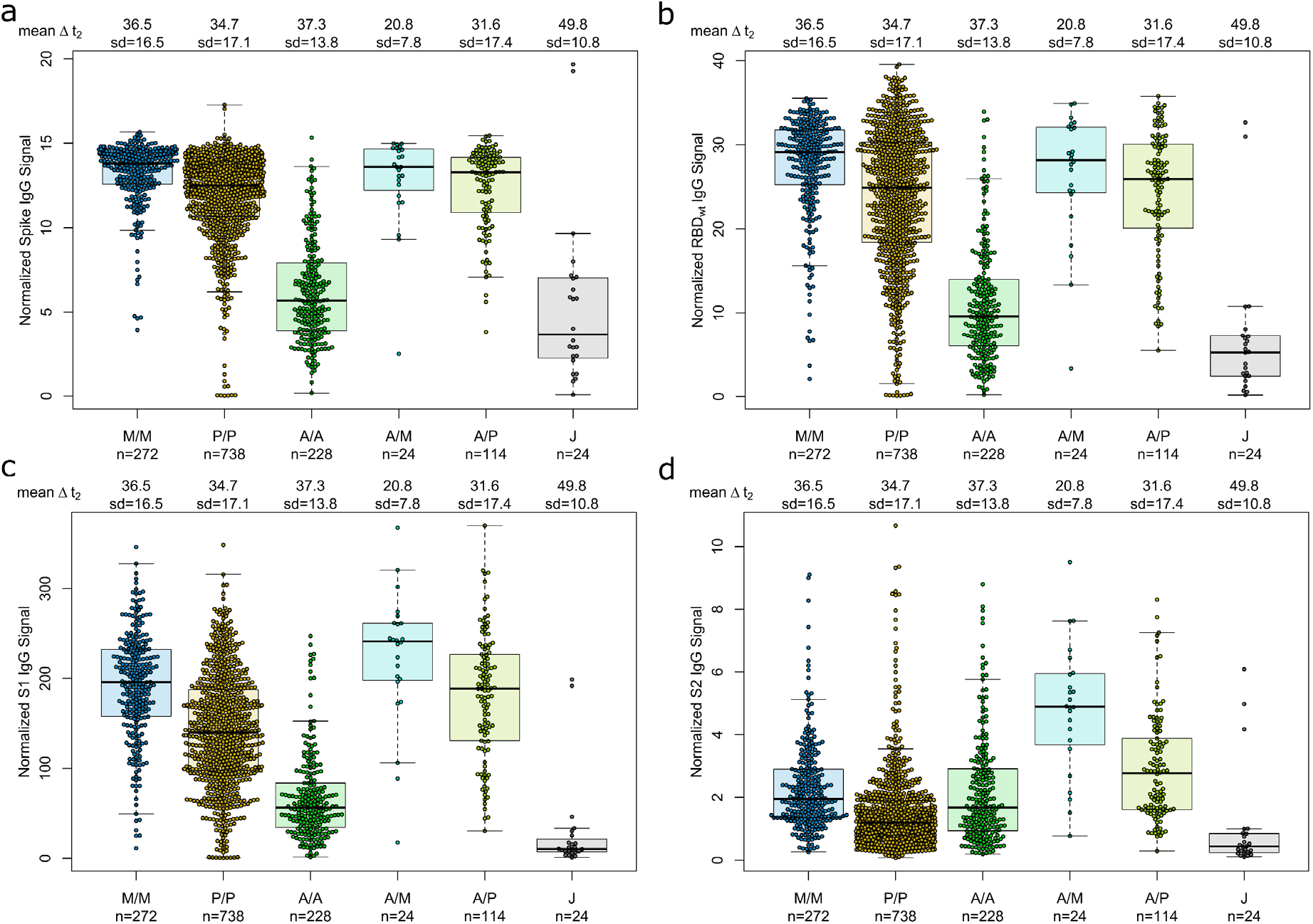
Different SARS-CoV-2 vaccination schemes result in distinct humoral responses. IgG antibody titres against full-length Spike trimer (a), receptor-binding domain (RBD) (b), S1 domain (c) and S2 domain (d) were measured with MULTICOV-AB. Individuals received either homologous mRNA-1273 (M/M, blue, n=272), BNT162b2 (P/P, orange, n=738) or AZD1222 (A/A, green, n=228), heterologous AZD1222-mRNA-1273 (A/M, light blue, n=24), AZD1222-BNT162b2 (A/P, light green, n=114), or a single dose of Ad26.CoV2.S (J, grey, n=24). Raw MFI values were normalised against QC samples to generate signal ratios for each antigen. Data is shown as box and whisker plots overlaid with strip charts. Boxes represent medians, 25th and 75th percentiles and whiskers show the largest and smallest non-outlier values based on 1.5 IQR calculation. Time between sampling and full vaccination is displayed as mean and SD for each group. Number of samples per vaccination scheme are stated below.

As multiplex-based serology tests such as MULTICOV-AB offer the unique opportunity for in-depth profiling of polyclonal antibody reactivity towards multiple viral antigens, we then assessed differences in antibody specificities between the different vaccines. Within the mix and match sample cohort, we observed that mRNA-based SARS-CoV-2 vaccinations resulted in reduced S2-specific antibody titres compared to vector-based ones **(Fig. 1)**. To investigate this unequal antibody distribution further, we initially scaled titres for each individual antigen **(Fig. 2)**, and found that while Spike, RBD, S1 titres were low for both AZD1222 and Ad26.CoV2.S, S2-specific titres were considerably higher than expected. We then calculated proportional ratios between antigens **(Table 3)**, confirming that homologous mRNA vaccination resulted in significantly higher proportion of RBD-(mRNA-1273 14.01-fold, BNT162b2 18.63-fold, AZD1222 5.23-fold) and S1-targeted antibodies (mRNA-1273 97.21-fold, BNT162b2 110.10-fold, AZD1222 33.48-fold) compared to S2-targeted immunoglobulins. This over-representation of S1-targeting antibodies following mRNA vaccination, was also present in those who received a heterologous dose schedule (AZD1222-mRNA-1273 47.60-fold, AZD1222-BNT162b2 65.06-fold).

**Table 3.**
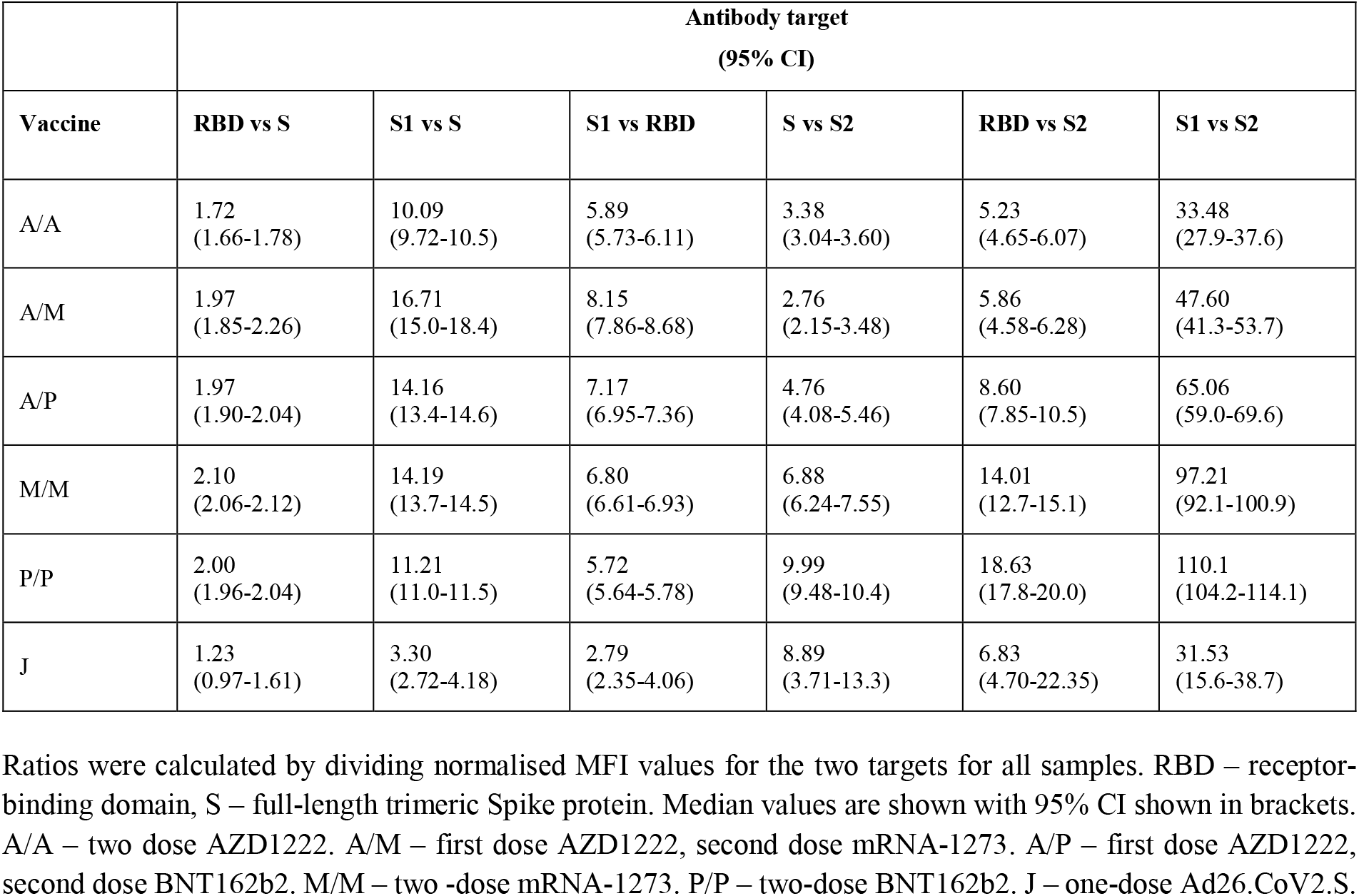
Antigen-specific ratios for different vaccination schemes.

**Fig. 2.**
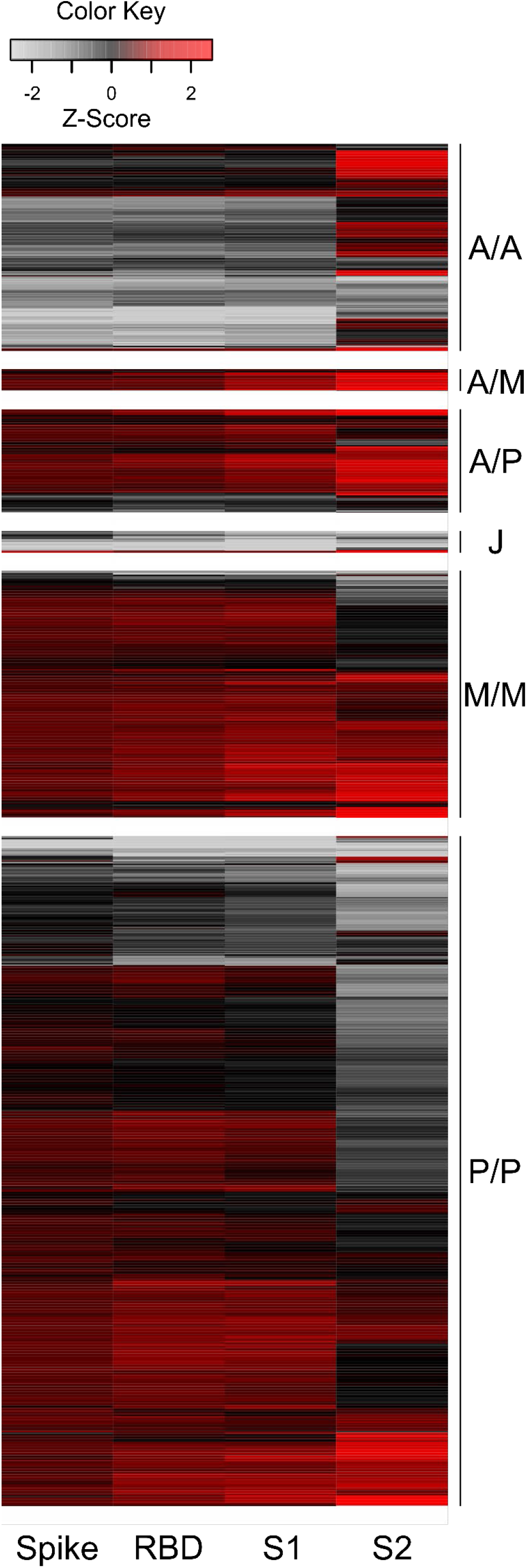
Humoral immune response after mRNA vaccination is skewed towards increased RBD and S1 titres, while vector-based vaccination results in increased S2 antibody levels. Antigen-specific antibody titres measured in the mix and match sample cohort were scaled and centred per antigen. Resulting values greater than 2.5 and smaller than −2.5 were set to these extreme values instead. Samples were then clustered within their subgroups based on immunisation scheme and are displayed as a heat map. Negative values represent below average titres and positive values represent positive above average titres per antigen. Colour shades indicate low (grey) to high (red) value distribution. A/A – two-dose AZD1222. A/M – first dose AZD1222, second dose mRNA-1273. A/P – first dose AZD1222, second dose BNT162b2. M/M – two-dose mRNA-1273. P/P – two-dose BNT162b2. J – one-dose Ad26.CoV2.S.

Having determined that mRNA-vaccines produce a significantly higher proportion of RBD and S1 antibodies, we next investigated their ACE2 binding inhibition as these antigens are predominantly responsible for antibody-mediated virus neutralization (12, 13). For this, we used a previously published RBD-ACE2 competition assay (21, 29), which detects neutralizing antibody activity only and is comparable to classical viral neutralization assays (9, 29). As expected, homologous mRNA vaccination resulted in higher ACE2 binding inhibition than homologous vector-based vaccination (median ACE2 binding inhibition mRNA-1273 0.93, BNT162b2 0.80, AZD1222 0.39, Ad26.CoV2.S 0.03, **Fig. 3)**.

**Fig. 3.**
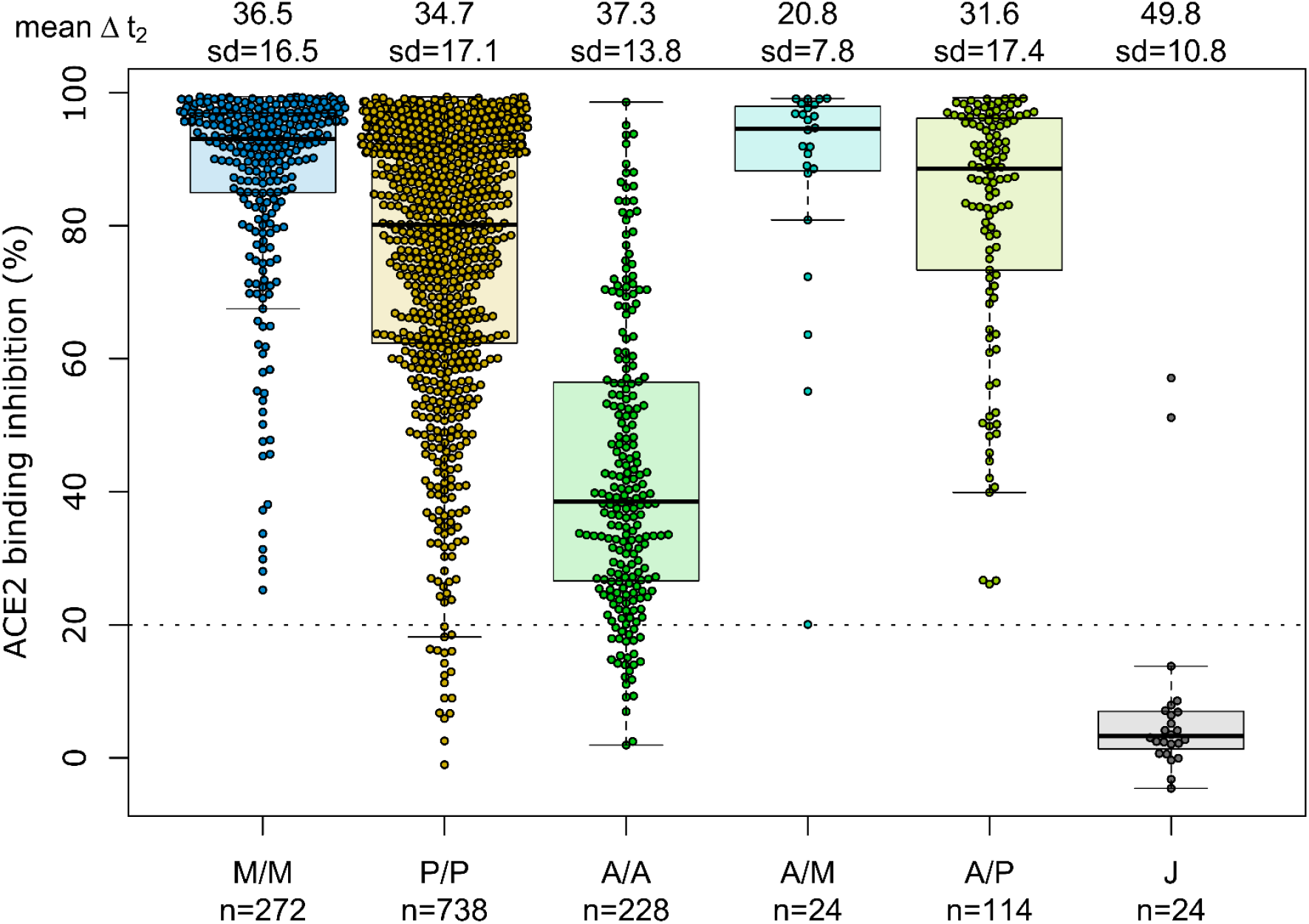
Different vaccination schemes impact ACE2 binding inhibition against SARS-CoV-2 wild-type. ACE2 binding inhibition against the SARS-CoV-2 wild-type (B1 isolate) RBD was assessed by an ACE2-RBD competition assay for homologous mRNA (mRNA-1273 (M/M, blue), BNT162b2 (P/P, orange)), heterologous prime-boost (AZD1222-mRNA-1273 (A/M, light blue), AZD1222-BNT162b2 (A/P, light green) or vector-based (AZD1222-AZD1222 (A/A, green), Ad26.CoV2.S (J, grey)) vaccination schemes in the mix and match cohort. Data is shown as box and whisker plots overlaid with strip charts. Boxes represent medians, 25th and 75th percentiles and whiskers show the largest and smallest non-outlier values based on 1.5 IQR calculation. The threshold for non-responsive samples (ACE2 binding inhibition less than 20%) is shown as dotted line. All samples below this threshold can be considered non-responsive. Time between sampling and full vaccination is displayed as mean and SD for each group. Number of samples per vaccination scheme are stated below. ACE2 binding inhibition towards VoCs can be found in Fig. S2.

Neutralizing antibodies generated following vaccination with Ad26.CoV2.S resulted in minimal ACE2 binding inhibition, with only 8.3% being classified as responders (29). As variants of concern now comprise the majority of infections globally (36), we also assessed ACE2 binding inhibition against the Alpha, Beta, Gamma and Delta SARS-CoV-2 VoC strains. ACE2 binding inhibition was most similar to wild-type for the Alpha variant, followed by Delta whereas Beta and Gamma variants had the largest reductions in ACE2 binding inhibition (**Fig. S2**).

Due to the range of responses recorded for each dose combination and likely differences in population characteristics as a result of changing vaccine recommendations, we examined whether confounders (sampling time post-vaccination (ΔT), age, gender or comorbidities) were instead responsible. To analyse impact of ΔT, we separated samples into 7 to 27 days post-final dose to capture peak response and 28 to 65 days post-final dose to capture plateau response (**Fig. 4)**. While there was a reduction in median response for samples from individuals collected within the plateau phase, the pattern between the vaccines remained consistent. While increasing age did result in small reductions in ACE2 binding inhibition (only significant for BNT162b2, p<0.001), the vaccine dosing scheme received had a substantially larger effect, with the eldest age group (>79) of homologous mRNA vaccine recipients still having increased IgG titres and ACE2 inhibition capacities than the youngest (26 to 45) AZD1222 recipients (**Fig. 4**). Regression modelling for ACE2 binding inhibition against wild-type confirmed the decrease of ACE2 binding inhibition with time post-vaccination for all vaccination types except homologous AZD1222 (**Table S4A**). While age did not cause a significant decrease for homologous AZD1222, this may have been due to the low number of samples at both ends of the age range within our cohort. For mRNA-1273, while age did result in a significant decrease during the peak period (p=0.029), this was not present within the plateau phase (p=0.615). For homologous BNT162b2 vaccination, male sex seemed to be associated with a decreased ACE2 binding inhibition, although the same was not true for mRNA-1273. Similar patterns were observed for the ACE2 binding inhibition against Alpha, Beta, Gamma and Delta VoCs (**Table S4B-E**). As we observed serological non-responders within our mix and match study cohort, we systematically evaluated their distribution among the different immunisation schemes (**Table 2**). Overall, vector-based homologous vaccination (2.78%) resulted in a higher proportion of non-responders than homologous mRNA-based vaccination (0.89%). Neither age nor gender was a determining factor in being a non-responder.

**Fig. 4.**
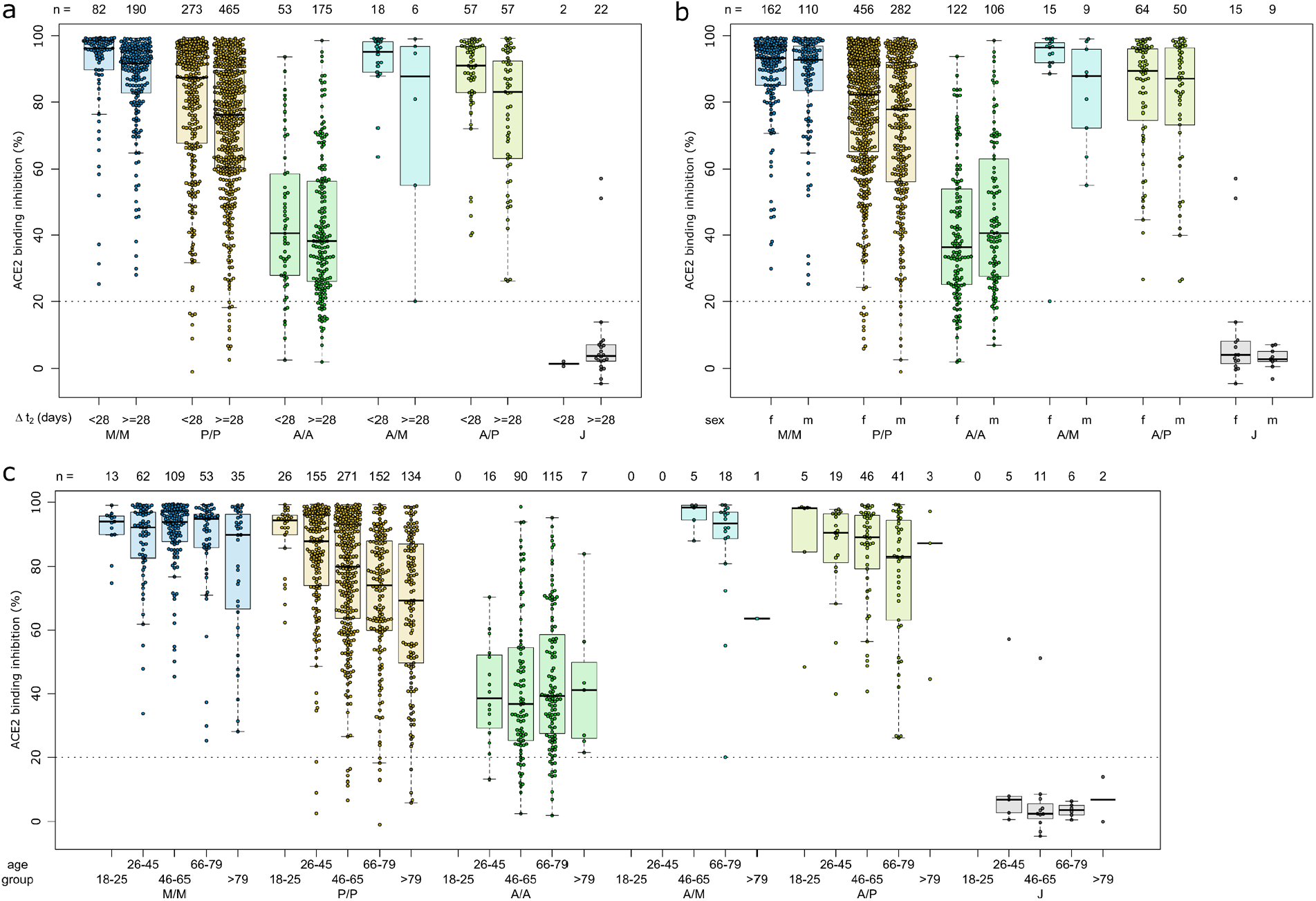
Effect of time post-vaccination, gender and age on ACE2 binding inhibition for different SARS-CoV-2 vaccination schemes. ACE2 binding inhibition against the SARS-CoV-2 wild-type (B1 isolate) RBD was assessed by an ACE2-RBD competition assay. Samples were split according to vaccination scheme (homologous mRNA (mRNA-1273 (M/M, blue), BNT162b2 (P/P, orange)), heterologous prime-boost (AZD1222-mRNA-1273 (A/M, light blue), AZD1222-BNT162b2 (A/P, light green) or vector-based (AZD1222-AZD1222 (A/A, green), Ad26.CoV2.S (J, grey)). To display impact of potential confounders, samples were further split in time post-vaccination up to 27 and above 27 days (a), gender (b) and indicated age groups (c). Boxes represent medians, 25th and 75th percentiles and whiskers show the largest and smallest non-outlier values based on 1.5 IQR calculation. The threshold for non-responsive samples (ACE2 binding inhibition less than 20%) is shown as dotted line. All samples below this threshold can be considered non-responsive. Time between sampling and full vaccination is displayed as mean and SD for each group. Number of samples per vaccination scheme are stated below the figure. Statistical significance was calculated by a regression model (Table S4).

As our population-based cohort also contained individuals who had been previously infected and then vaccinated, we examined what effect this had upon their vaccine-induced response. As previously observed (37), recovered and then vaccinated individuals developed high levels of IgG with strong ACE2 binding inhibition (**Fig. 5, Table S6**). This increase was particularly apparent for the vector-based vaccinations where median RBD IgG titres (AZD1222 24.69, Ad26.CoV2.S 36.53, **Fig. S3**) and median ACE2 binding inhibition (AZD1222 0.93, Ad26.CoV2.S 0.71, **Fig. 5**) were significantly higher than in SARS-CoV-2 naïve vaccinated individuals (median RBD IgG AZD1222 9.61, Ad26.CoV2.S 5.25, **Fig. 1**) and median ACE2 binding inhibition (AZD1222 39%, Ad26.CoV2.S 0.03%, **Fig. 3**).

**Fig. 5.**
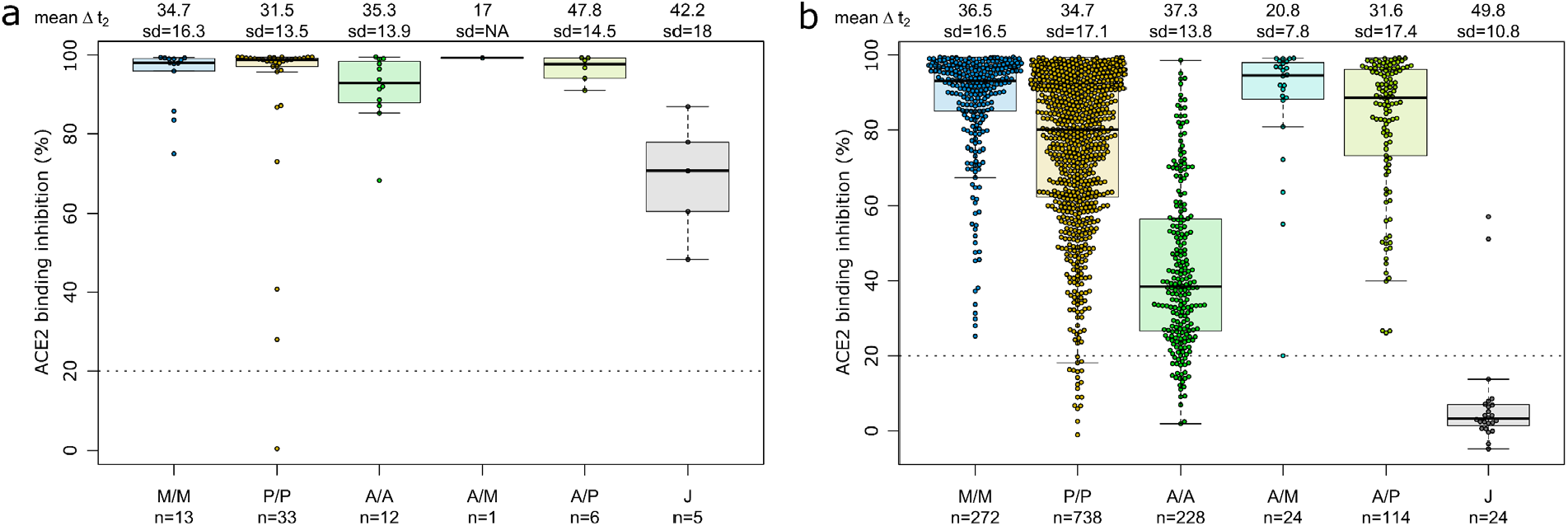
SARS-CoV-2 vaccination boosts ACE2 binding inhibition among recovered individuals independent of vaccination scheme. Differences in vaccination responses of recovered previously SARS-CoV-2 infected individuals from our mix and match cohort were analysed using an ACE2-RBD competition assay (a). SARS-CoV-2 infection status was based on a previous self-reported positive PCR/antigen test or a MULTICOV-AB nucleocapsid IgG normalisation ratio above 1. Samples were split according to vaccination scheme (homologous mRNA (mRNA-1273 (M/M, blue), BNT162b2 (P/P, orange)), heterologous prime-boost (AZD1222-mRNA-1273 (A/M, light blue), AZD1222-BNT162b2 (A/P, light green) or vector-based (AZD1222-AZD1222 (A/A, green), AdCoV2S (J, grey)). For clarity and comparison, ACE2 inhibition of SARS-CoV-2 naïve individuals are displayed (b). Boxes represent medians, 25th and 75th percentiles and whiskers show the largest and smallest non-outlier values based on 1.5 IQR calculation. If not enough sample to generate a box were present (minimum 5), then only the median is indicated by a line. The threshold for non-responsive samples (ACE2 binding inhibition less than 20%) is shown as dotted line. All samples below this can be considered non-responsive. Time between sampling and full vaccination is displayed as mean and SD for each group. Number of samples per vaccination scheme are stated below the figure. Results of a formal statistical comparison of recovered-vaccinated and SARS-CoV-2 naïve-vaccinated individuals are shown in Table S6.

Having determined that mRNA-based vaccination resulted in an increased humoral response, we evaluated lifespan and antibody response kinetics using our time point sample cohort which were selected to mimic key response periods for antibody-producing B-cell activity such as expansion, peak and plateau phase after a complete vaccination scheme. Vaccine-induced titres and ACE2 binding inhibition both initially increased, peaked during the second time point (26 to 30 days post-second dose), and then decreased linearly as time increased (**Fig. 6**). ACE2 binding inhibition followed the same pattern of decrease as time increased. In contrast to antibody levels, the percentage of non-responders showed however a trend for increased decline already from time point 94 to 103 days post-second vaccination onwards for BNT162b2, with 22.22% of samples considered as non-responders at 176-203 days post-second vaccination (**Table 2**). As already observed in Figure 1, mRNA-1273 (blue line) resulted in higher titres and ACE2 binding inhibition compared to BNT162b2 (yellow line) for all monitored time points. To validate this pattern of decreasing antibody titres and ACE2 inhibition activity, we examined samples from a cohort of longitudinal donors (longitudinal sample cohort). Unlike the time point sample cohort, this cohort contained paired samples from each donor which allows to directly compare changes in titre and activity from the first sampling to the second sampling. While these samples had a variable initial ΔT post-full vaccination (7-63 days), the sampling intervals between first and second donation were more comparable (114-163 days). Overall, titres decreased (median RBD 68%) between their first and second sampling (**Fig. 7**). Among the different SARS-CoV-2 antigens, RBD and S1 antibodies had the largest decrease, while Spike Trimer and S2 had the smallest. This reduction in titre was reflected in ACE2 binding inhibition which also reduced substantially from the first to second sampling (median 32%). ACE2 binding inhibition was also clearly decreased for all VoC RBDs (Alpha 36%, Beta 30%, Gamma 29%, Delta 38%).

**Fig. 6.**
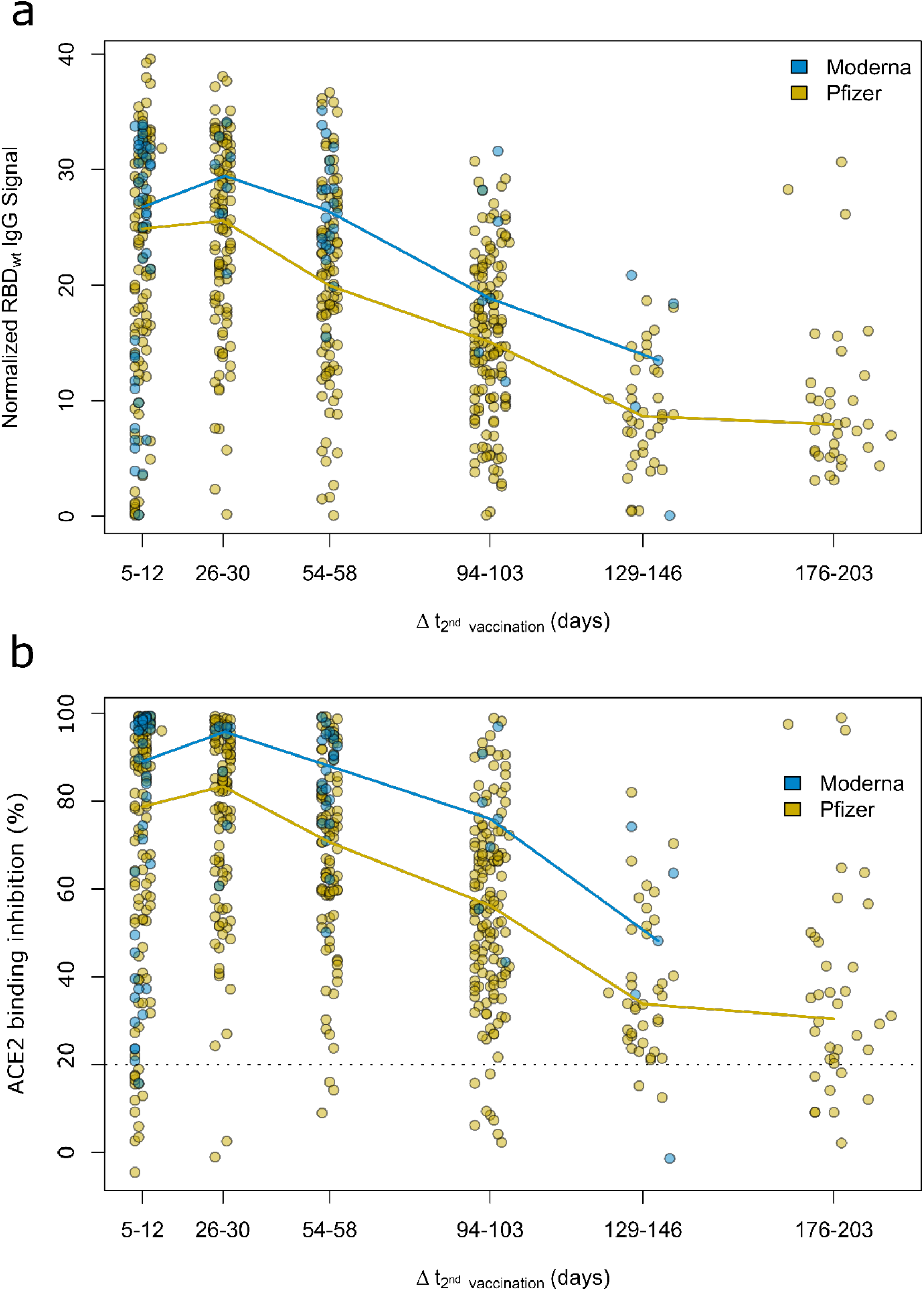
Antibody and neutralization response kinetic up to 7 months after SARS-CoV-2 mRNA–vaccination. Humoral vaccine response was assessed by MULTICOV-AB (a) and an ACE2-RBD competition assay (b) using the time point sample set. Samples were either 5 to 12, 26 to 30, 54 to 58, 94 to 103, 129 to 146 and 176 to 203 days post-second dose of either a two-dose BNT162b2 (yellow, n=515) or mRNA-1273 (blue, n=82) vaccination. Coloured line connects median response per time point and vaccine. Data is displayed as normalised IgG RBD ratio or as ACE2 binding inhibition % where 100% indicates maximum binding inhibition and 0% no binding inhibition. The threshold for non-responsive samples (ACE2 binding inhibition less than 20%) is shown as dotted line. All samples below this threshold can be considered non-responsive.

**Fig. 7.**
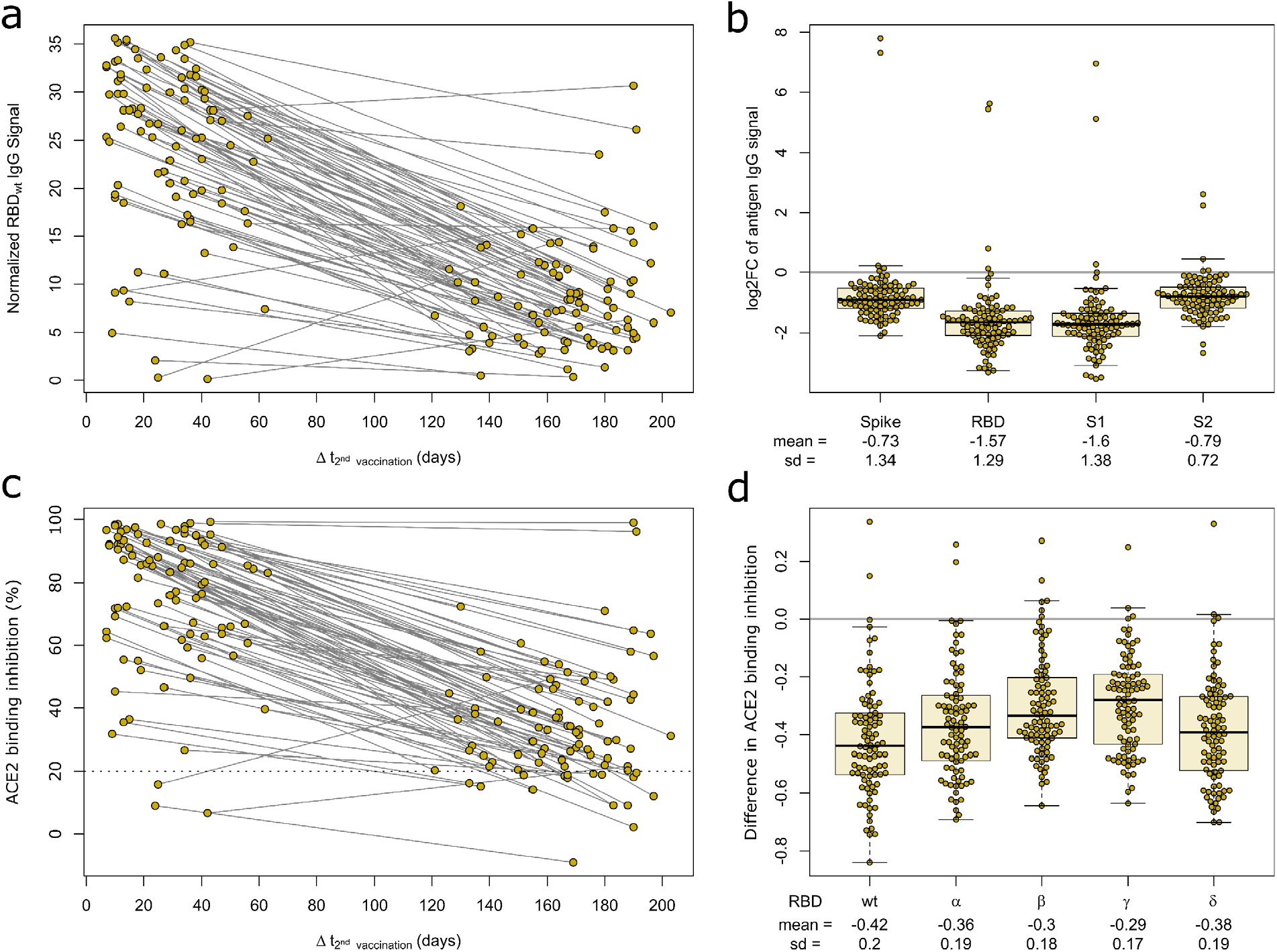
Longitudinal mRNA vaccine response monitoring defines decreases over time. Humoral response was assessed by MULTICOV-AB (a, b) and an ACE2-RBD competition assay (c, d) for all samples of the longitudinal sample set at indicated time points post-second dose of a complete BNT162b2 vaccination scheme (n=90). Line graph illustrates longitudinal development of RBD-specific antibody levels (a) or ACE2 binding inhibition (c) towards WT RBD as time post-vaccination increases. The threshold (less than 20% ACE2 binding inhibition) for non-responsive samples within the ACE2-RBD competition assay is shown (dotted line). All samples below this threshold are considered non-responsive. SARS-CoV-2 antigen-specific differences in longitudinal humoral response are expressed as log2 fold changes (b). Reduction in ACE2 binding inhibition for RBD of Alpha, Beta, Gamma and Delta VoC between sampling times is shown as difference (d).

## 4. Discussion

We report both significant and substantial differences in humoral responses generated by the different vaccines and dose regimens currently available in Germany, with homologous mRNA or combined heterologous vector and mRNA vaccination approaches inducing significantly higher titres and ACE2 binding inhibition compared to homologous vector-based vaccination schemes. This expands on results from on-going randomised and observational trials such as the ComCoV (38) or CoCo (39) study which provided only information on AZD1222-BNT162b2 schemes (19, 40). Among homologous mRNA regimens, we identified like others, that mRNA-1273 resulted in higher antibody titres and ACE2 binding inhibition than BNT162b2 (37, 41). While we have used an ACE2-RBD competition assay to measure ACE2 binding inhibition as opposed to classic virus neutralization assays, this assay analyses neutralizing antibodies as seen by its similar performance to VNT (9, 29). ACE2 inhibition assays instead of a VNT have also already been used successfully by other groups to determine neutralizing antibody activity (42). Methodically, MULTICOV-AB and the ACE2-RBD competition assay are also complementary and are measured using a single initial sample dilution which further reduces variability between their results. As expected, titres and ACE2 binding inhibition for AZD1222 were reduced compared to mRNA-based vaccination (40). By multiplex-based antibody profiling, we were able to investigate these differences and determine that vector and mRNA-based vaccines induced a distinct pattern of Spike subdomain-targeted antibodies. While vector-based formulations result in a significantly larger proportion of S2-domain antibodies, RBD- and S1-domain antibodies dominated in mRNA vaccines. While these observations require further detailed investigation, the relative over-representation of RBD- and S1-targeting antibodies within mRNA vaccines is particularly intriguing as these two antigens comprise the majority of neutralizing antibody activity (13). Despite the lack of a clearly defined correlation of vaccine efficacy and neutralizing antibody levels, it appears logical that increased antibody levels specific to virus protein-mediating cell attachment could result in enhanced levels of protection from infection and contribute to observed differences in levels of vaccine efficacy and effectiveness (2, 43, 44). Interestingly, a series of modelling studies have now linked levels of neutralizing antibodies to vaccine efficacy (12, 45).

An additional finding of our study requiring further investigation is the relatively poor performance of Ad26.CoV2.S, particularly for induction of neutralizing antibodies for both SARS-CoV-2 wild-type and VoC RBDs. While some studies have reported sufficient levels of neutralizing activity after vaccination with Ad26.CoV2.S (4), others identified minimal neutralizing activity, particularly when compared to other COVID-19 vaccines from Pfizer or Moderna (41). The relatively poor performance of Ad26.CoV2.S in inducing an antibody response has also been identified by researchers studying other bodily fluids (e.g. breast milk), who found that Ad26.CoV2.S produced significantly fewer IgA antibodies than BNT162b2 or mRNA-1273 (46). While our Ad26.CoV2.S sample group size is low (n=29), it is three times larger than a recent study from the manufacturer which reported neutralizing activity against Delta and other VoCs (n=8, (47)). It should be noted that four of the eight individuals within their cohort were reported as being spike seropositive at baseline which is a consistent finding with our cohort, where strong ACE2 binding inhibition was only achieved in those individuals who had been previously infected. Our median time point is however earlier than the reported peak of antibody activity (4, 48). Further independent investigations into the neutralizing activity generated by single-dose of Ad26.CoV2.S to clarify those differing results within SARS-CoV-2 naïve individuals are therefore urgently needed.

Among confounding variables, we identified like others that age resulted in a general reduction in titre and ACE2 binding inhibition (10, 37, 49), although the vaccine dose scheme received had a more significant effect. While recovered individuals developing high titres and ACE2 binding inhibition once vaccinated has been previously reported (37, 40), we found that these responses were similar among all vaccines and dose schedules. Given that current German guidelines require a six month post-positive PCR waiting period before receiving a first dose, this suggests that such individuals would be suitable for all currently licensed vaccines, assuming they meet pre-existing EMA and STIKO criteria. This ability to use all vaccines and generate a substantial response will be of particular public health importance, given the on-going booster dose administration which could impact availability for some vaccine brands, as happened earlier in 2021.

Our results on the longevity of the humoral response post-vaccination is similar to others, in identifying an initial peak from approximately 28 days post-second dose onwards followed by a gradual reduction over time (50). As expected, ACE2 binding inhibition and titre are mostly mirrored in their decline over time. However, the increased numbers of non-responders from BNT162b2-vaccinated individuals from six months after the second vaccine needs further careful monitoring until a precise correlate of protection has been defined. Among the different SARS-CoV-2 antibodies, it is unsurprising that the RBD and S1 underwent the greatest reductions as they had the largest titres to begin with. Between VoCs, we did not identify any apparent differences in ACE2 binding inhibition within the differing vaccines and dose regimens for confounders. Instead, again vaccine or dose regimen received had the largest effect upon ACE2 binding inhibition. The VoCs themselves followed a previously published pattern (9, 21, 51), with the lowest reduction for the Alpha variant, and the highest for the Beta and Gamma variants. It should be stated that we in our analysis of longitudinal samples, there is a wide variety of timeframes post-vaccination, meaning that initial samples are collected both before, during and after the initial peak response at 28 days. While we have then made the assumption that decreases in responses would be linear to the second sampling, this is not the case as some of the early collected samples (e.g. 7 days post-second vaccination) would have initially increased before later decreasing. However, our purpose of this analysis was to measure changes over a larger timeframe (4 months) and the difference in time from first to second sampling, means that all samples should be in the decline phase by their second sampling.

Our manuscript has several limitations, namely that we are only measuring antibodies (including neutralizing antibodies) that are present within serum. As previously stated, we have used an ACE2-RBD competition assay to measure inhibition of ACE2 binding instead of classical virus neutralization assays, although the results of this assay have already been shown to be similar to VNT and are known to be specific to neutralizing antibody responses only. While neutralizing antibodies themselves are considered a strong correlate for protection (13), other components that are not measured within our assays such as T-cell mediated immunity will also offer protection (52, 53). Our use of serum also means that memory B-cells, which are involved in protection against severe disease progression (54), are equally excluded from our analysis. Our study cohort consists of relatively low sample numbers for both heterologous and Ad26.CoV2.S vaccinations whereas BNT162b2 samples are overrepresented. However, our sample numbers are similar or in the case of Ad26.CoV2.S exceed other previously published work making our study one of the largest independent evaluation studies of this vaccine. Our BNT162b2 sample size mimics dose distribution in Germany where approximately 70% of delivered vaccine doses were from Pfizer. Our study population is also relatively similar in regard to age and gender.

Overall, we provide data on the vaccine-induced humoral response for all currently available mRNA-, vector-based and heterologous immunisation regimens in Germany. Within our population-based cohort, mRNA homologous or heterologous vaccination resulted in increased humoral responses. Our multiplex approach identified differences in quantities and ratios of RBD- and S1-targeting antibodies following mRNA homologous or heterologous vaccination. Further investigation into this targeting will be of particular interest to improve vaccine performance particularly for next generation vector-based vaccines.

## Supporting information

Supplementary Information Comparative magnitude and persistence of SARS-CoV-2 vaccination responses on a population level in Germany

## Data Availability

Data is available upon request from the corresponding authors.

## Contributors

MS, AD, BL, GK and NSM conceived the study. NSM, MS, BL, VM, SC, and GK procured funding. AD, MS, MB and NSM, designed the experiments. AD, JG, JJ, and DJ performed the experiments. AD, MS, MB, MH, JO, SC, NW, SG, JH and BK performed data collection and analysis. AD, MB, MH, BK generated figures and tables. TT, KF, TI and NK organized sample collection and processing. AD, MS, MH and BK verified the underlying data. AD, MS and MB wrote the manuscript. AD, MH, JO, PH, BL, NSM, MS, YK, DG, VM, DJ, PDK, BT, UR, TT, KF, NK, TI, TK, AR, CS, AR, AM, NG and contributed resources or were involved in project administration. All authors critically reviewed and approved the final manuscript.

## Acknowledgments

First and foremost, we would like to thank the MuSPAD participants for their willingness and commitment to make this study possible. We also want to thank all laboratory members and administrative staff at the Institute of Transfusion Medicine and Immunohematology in Plauen for continued excellent technical and organizational support in sample processing. We are grateful to the entire team of BOS112 for running of MuSPAD study sites. We sincerely thank all participating counties, cities and local health care authorities for their support. We thank Astrid Hans and Carina Lützow for administrative assistance. We thank members of the Multiplex Immunoassays Group at the NMI for their assistance in sample arrival and storage.

## Data Sharing Statement

Data is available upon request from the corresponding authors.

## References

1. Voysey M, Clemens SAC, Madhi SA, Weckx LY, Folegatti PM, Aley PK, et al. Safety and efficacy of the ChAdOx1 nCoV-19 vaccine (AZD1222) against SARS-CoV-2: an interim analysis of four randomised controlled trials in Brazil, South Africa, and the UK. The Lancet. 2021;397(10269):99–111.

2. Baden LR, El Sahly HM, Essink B, Kotloff K, Frey S, Novak R, et al. Efficacy and Safety of the mRNA-1273 SARS-CoV-2 Vaccine. N Engl J Med. 2021;384(5):403–16.

3. Polack FP, Thomas SJ, Kitchin N, Absalon J, Gurtman A, Lockhart S, et al. Safety and Efficacy of the BNT162b2 mRNA Covid-19 Vaccine. New England Journal of Medicine. 2020;383(27):2603–15.

4. Sadoff J, Gray G, Vandebosch A, Cárdenas V, Shukarev G, Grinsztejn B, et al. Safety and Efficacy of Single-Dose Ad26.COV2.S Vaccine against Covid-19. New England Journal of Medicine. 2021;384(23):2187–201.

5. RKI. COVID-19 Dashboard https://experience.arcgis.com/experience/478220a4c454480e823b17327b2bf1d4/page/page_1/2021 [

6. Corona.stat.uni-muenchen.de/maps/. https://corona.stat.uni-muenchen.de/maps/2021 [

7. Scobie HM, Johnson AG, Suthar AB, Severson R, Alden NB, Balter S, et al. Monitoring Incidence of COVID-19 Cases, Hospitalizations, and Deaths, by Vaccination Status - 13 U.S. Jurisdictions, April 4-July 17, 2021. MMWR Morbidity and mortality weekly report. 2021;70(37):1284–90.

8. Government I. Verified results after receiving the vaccine among the first vaccinated group. In: Committee VEaSFU, editor. https://www.gov.il/BlobFolder/reports/vaccine-efficacy-safety-follow-up-committee/he/files_publications_corona_two-dose-vaccination-data.pdf2021.

9. Dulovic A, Strengert M, Ramos GM, Becker M, Griesbaum J, Junker D, et al. Diminishing immune responses against variants of concern in dialysis patients four months after SARS-CoV-2 mRNA vaccination. medRxiv. 2021:2021.08.16.21262115.

10. Naaber P, Tserel L, Kangro K, Sepp E, Jürjenson V, Adamson A, et al. Dynamics of antibody response to BNT162b2 vaccine after six months: a longitudinal prospective study. The Lancet Regional Health – Europe.

11. Kustin T, Harel N, Finkel U, Perchik S, Harari S, Tahor M, et al. Evidence for increased breakthrough rates of SARS-CoV-2 variants of concern in BNT162b2-mRNA-vaccinated individuals. Nature Medicine. 2021;27(8):1379–84.

12. Earle KA, Ambrosino DM, Fiore-Gartland A, Goldblatt D, Gilbert PB, Siber GR, et al. Evidence for antibody as a protective correlate for COVID-19 vaccines. Vaccine. 2021;39(32):4423–8.

13. Khoury DS, Cromer D, Reynaldi A, Schlub TE, Wheatley AK, Juno JA, et al. Neutralizing antibody levels are highly predictive of immune protection from symptomatic SARS-CoV-2 infection. Nature Medicine. 2021;27(7):1205–11.

14. Althaus K, Möller P, Uzun G, Singh A, Beck A, Bettag M, et al. Antibody-mediated procoagulant platelets in SARS-CoV-2-vaccination associated immune thrombotic thrombocytopenia. Haematologica. 2021;106(8):2170–9.

15. Greinacher A, Thiele T, Warkentin TE, Weisser K, Kyrle PA, Eichinger S. Thrombotic Thrombocytopenia after ChAdOx1 nCov-19 Vaccination. New England Journal of Medicine. 2021;384(22):2092–101.

16. Pottegård A, Lund LC, Karlstad Ø, Dahl J, Andersen M, Hallas J, et al. Arterial events, venous thromboembolism, thrombocytopenia, and bleeding after vaccination with Oxford-AstraZeneca ChAdOx1-S in Denmark and Norway: population based cohort study. BMJ. 2021;373:n1114.

17. Wise J. Covid-19: European countries suspend use of Oxford-AstraZeneca vaccine after reports of blood clots. BMJ. 2021;372:n699.

18. Vygen-Bonnet S, Koch J, Bogdan C, Harder T, Heininger U, Kling K, et al. Beschluss der STIKO zur 5. Aktualisierung der COVID-19-Impfempfehlung und die dazugehörige wissenschaftliche Begründung. 2021(19):24–36.

19. Barros-Martins J, Hammerschmidt SI, Cossmann A, Odak I, Stankov MV, Morillas Ramos G, et al. Immune responses against SARS-CoV-2 variants after heterologous and homologous ChAdOx1 nCoV-19/BNT162b2 vaccination. Nature Medicine. 2021;27(9):1525–9.

20. Schmidt T, Klemis V, Schub D, Mihm J, Hielscher F, Marx S, et al. Immunogenicity and reactogenicity of heterologous ChAdOx1 nCoV-19/mRNA vaccination. Nature Medicine. 2021;27(9):1530–5.

21. Strengert M, Becker M, Ramos GM, Dulovic A, Gruber J, Juengling J, et al. Cellular and humoral immunogenicity of a SARS-CoV-2 mRNA vaccine in patients on haemodialysis. EBioMedicine. 2021;70.

22. Carr EJ, Wu M, Harvey R, Wall EC, Kelly G, Hussain S, et al. Neutralising antibodies after COVID-19 vaccination in UK haemodialysis patients. The Lancet. 2021;398(10305):1038–41.

23. Harsch IA, Ortloff A, Reinhöfer M, Epstude J. Symptoms, antibody levels and vaccination attitude after asymptomatic to moderate COVID-19 infection in 200 healthcare workers. GMS hygiene and infection control. 2021;16:Doc15.

24. Becker M, Dulovic A, Junker D, Ruetalo N, Kaiser PD, Pinilla YT, et al. Immune response to SARS-CoV-2 variants of concern in vaccinated individuals. Nature Communications. 2021;12(1):3109.

25. Galanis P, Vraka I, Fragkou D, Bilali A, Kaitelidou D. Seroprevalence of SARS-CoV-2 antibodies and associated factors in healthcare workers: a systematic review and meta-analysis. The Journal of hospital infection. 2021;108:120–34.

26. Tregoning JS, Flight KE, Higham SL, Wang Z, Pierce BF. Progress of the COVID-19 vaccine effort: viruses, vaccines and variants versus efficacy, effectiveness and escape. Nature Reviews Immunology. 2021;21(10):626–36.

27. Gornyk D, Harries M, Glöckner S, Strengert M, Kerrinnes T, Heise J-K, et al. SARS-CoV-2 seroprevalence in Germany. Dtsch Arztebl International. 2021;0(OnlineFirst):1-.

28. Becker M, Strengert M, Junker D, Kaiser PD, Kerrinnes T, Traenkle B, et al. Exploring beyond clinical routine SARS-CoV-2 serology using MultiCoV-Ab to evaluate endemic coronavirus cross-reactivity. Nature Communications. 2021;12(1):1152.

29. Junker D, Dulovic A, Becker M, Wagner TR, Kaiser PD, Traenkle B, et al. Reduced serum neutralization capacity against SARS-CoV-2 variants in a multiplex ACE2 RBD competition assay. medRxiv. 2021:2021.08.20.21262328.

30. Renk H, Dulovic A, Becker M, Fabricius D, Zernickel M, Junker D, et al. Typically asymptomatic but with robust antibody formation: Children’s unique humoral immune response to SARS-CoV-2. medRxiv. 2021:2021.07.20.21260863.

31. Planatscher H, Rimmele S, Michel G, Potz O, Joos T, Schneiderhan-Marra N. Systematic reference sample generation for multiplexed serological assays. Sci Rep. 2013;3.

32. R Core Team. R: A language and environment for statistical computing. Vienna, Austria: R Foundation for Statistical Computing; 2021.

33. Holm S. A Simple Sequentially Rejective Multiple Test Procedure. Scandinavian Journal of Statistics. 1979;6(2):65–70.

34. Warnes GR, Bolker B, Bonebakker L, Gentleman R, Huber W, Liaw A, et al. gplots: Various R Programming Tools for Plotting Data. 3.1.1 ed2021.

35. Eklund A, Trimble J. beeswarm: The Bee Swarm Plot, an Alternative to Stripchart. 0.4.0 ed2021.

36. CoVariants. Shared Mutations (https://covariants.org/shared-mutations) [

37. Steensels D, Pierlet N, Penders J, Mesotten D, Heylen L. Comparison of SARS-CoV-2 Antibody Response Following Vaccination With BNT162b2 and mRNA-1273. JAMA. 2021.

38. Liu X, Shaw RH, Stuart ASV, Greenland M, Aley PK, Andrews NJ, et al. Safety and immunogenicity of heterologous versus homologous prime-boost schedules with an adenoviral vectored and mRNA COVID-19 vaccine (Com-COV): a single-blind, randomised, non-inferiority trial. Lancet. 2021;398(10303):856–69.

39. Behrens GMN, Cossmann A, Stankov MV, Schulte B, Streeck H, Förster R, et al. Strategic Anti-SARS-CoV-2 Serology Testing in a Low Prevalence Setting: The COVID-19 Contact (CoCo) Study in Healthcare Professionals. Infectious Diseases and Therapy. 2020;9(4):837–49.

40. Eyre DW, Lumley SF, Wei J, Cox S, James T, Justice A, et al. Quantitative SARS-CoV-2 anti-spike responses to Pfizer-BioNTech and Oxford-AstraZeneca vaccines by previous infection status. Clin Microbiol Infect. 2021;27(10):1516.e7-.e14.

41. Tada T, Zhou H, Samanovic MI, Dcosta BM, Cornelius A, Mulligan MJ, et al. Comparison of Neutralizing Antibody Titers Elicited by mRNA and Adenoviral Vector Vaccine against SARS-CoV-2 Variants. bioRxiv. 2021:2021.07.19.452771.

42. Lopez E, Haycroft ER, Adair A, Mordant FL, O’Neill MT, Pymm P, et al. Simultaneous evaluation of antibodies that inhibit SARS-CoV-2 variants via multiplex assay. JCI Insight. 2021;6(16).

43. ChAdOx1 nCoV-19 Vaccine Efficacy against the B.1.351 Variant. New England Journal of Medicine. 2021;385(6):571–2.

44. Self WH, Tenforde MW, Rhoads JP, Gaglani M, Ginde AA, Douin DJ, et al. Comparative Effectiveness of Moderna, Pfizer-BioNTech, and Janssen (Johnson & Johnson) Vaccines in Preventing COVID-19 Hospitalizations Among Adults Without Immunocompromising Conditions-United States, March-August 2021. MMWR Morbidity and mortality weekly report. 2021;70(38):1337–43.

45. Feng S, Phillips DJ, White T, Sayal H, Aley PK, Bibi S, et al. Correlates of protection against symptomatic and asymptomatic SARS-CoV-2 infection. Nature Medicine. 2021.

46. Fox A, DeCarlo C, Yang X, Norris C, Powell RL. Comparative profiles of SARS-CoV-2 Spike-specific milk antibodies elicited by COVID-19 vaccines currently authorized in the USA. medRxiv. 2021:2021.07.19.21260794.

47. Jongeneelen M, Kaszas K, Veldman D, Huizingh J, van der Vlugt R, Schouten T, et al. Ad26.COV2.S elicited neutralizing activity against Delta and other SARS-CoV-2 variants of concern. bioRxiv. 2021:2021.07.01.450707.

48. Barouch DH, Stephenson KE, Sadoff J, Yu J, Chang A, Gebre M, et al. Durable Humoral and Cellular Immune Responses 8 Months after Ad26.COV2.S Vaccination. New England Journal of Medicine. 2021;385(10):951–3.

49. Shrotri M, Fragaszy E, Geismar C, Nguyen V, Beale S, Braithwaite I, et al. Spike-antibody responses to ChAdOx1 and BNT162b2 vaccines by demographic and clinical factors (Virus Watch study). medRxiv. 2021:2021.05.12.21257102.

50. Doria-Rose N, Suthar MS, Makowski M, O’Connell S, McDermott AB, Flach B, et al. Antibody Persistence through 6 Months after the Second Dose of mRNA-1273 Vaccine for Covid-19. New England Journal of Medicine. 2021;384(23):2259–61.

51. van Gils MJ, Lavell AHA, van der Straten K, Appelman B, Bontjer I, Poniman M, et al. Four SARS-CoV-2 vaccines induce quantitatively different antibody responses against SARS-CoV-2 variants. medRxiv. 2021:2021.09.27.21264163.

52. Mateus J, Dan Jennifer M, Zhang Z, Rydyznski Moderbacher C, Lammers M, Goodwin B, et al. Low-dose mRNA-1273 COVID-19 vaccine generates durable memory enhanced by cross-reactive T cells. Science.0(0):eabj9853.

53. Israelow B, Mao T, Klein J, Song E, Menasche B, Omer Saad B, et al. Adaptive immune determinants of viral clearance and protection in mouse models of SARS-CoV-2. Science Immunology.0(0):eabl4509.

54. Goel RR, Apostolidis SA, Painter MM, Mathew D, Pattekar A, Kuthuru O, et al. Distinct antibody and memory B cell responses in SARS-CoV-2 naïve and recovered individuals following mRNA vaccination. Sci Immunol. 2021;6(58).

