## Supplementary Information Comparative magnitude and persistence of SARS-CoV-2 vaccination responses on a population level in Germany for "Comparative magnitude and persistence of SARS-CoV-2 vaccination responses on a population level in Germany"

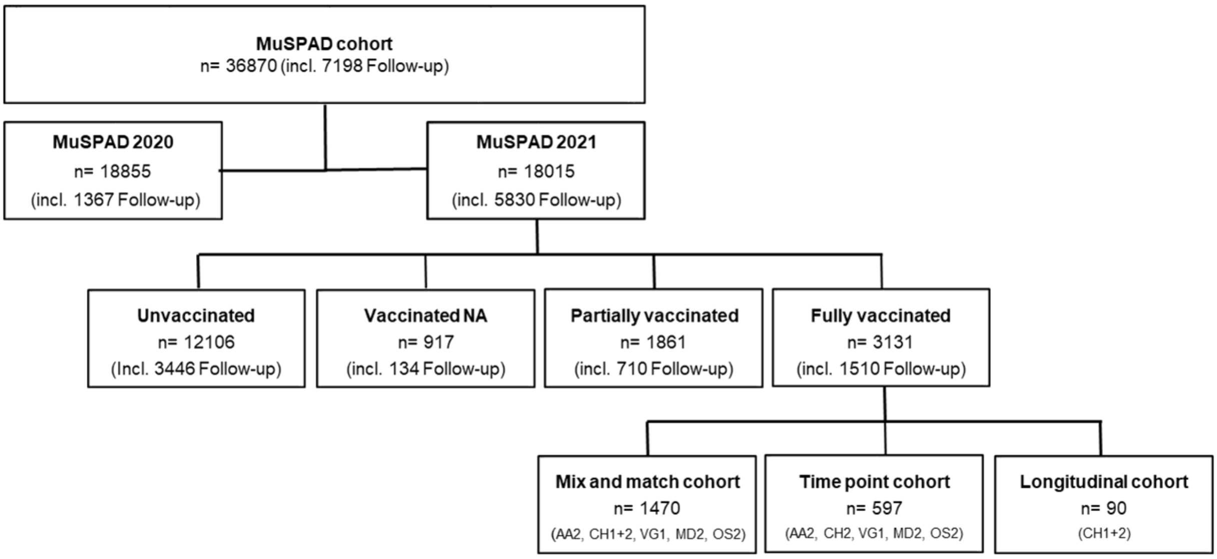


**Fig. S1. Flow chart for sample selection from MuSPAD cohort.**
Sample selection for our vaccine response study from the entire MuSPAD cohort is displayed in a flow chart. Samples were selected based on selection criteria outlined in method section to examine impact of vaccination scheme, length and persistence of antibody response. Samples originated from the following study locations: Aachen 2 (AA2), Chemnitz 1(CH1), Chemnitz 2 (CH2), Vorpommern-Greifswald 1 (VG1), Osnabrück 2 (OS2), and Magdeburg 2 (MD2). Number of participants is given per group (n), NA: not available.

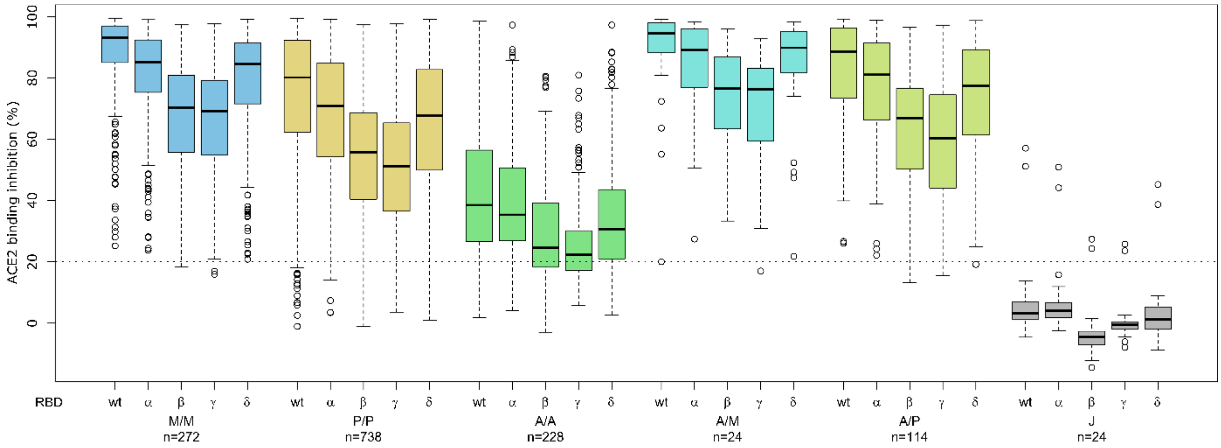


**Fig. S2. Different VoCs reduce antibody ACE2 binding inhibition comparably between SARS-CoV-2 vaccination schemes.**
ACE2 binding inhibition against the SARS-CoV-2 VoC Alpha (B.1.1.7), Beta (B.1.351), Gamma (P1) and Delta (B.1.617.2) RBDs was assessed by an ACE2-RBD competition assay for homologous mRNA (mRNA-1273 (M/M, blue), BNT162b2 (P/P, orange)), heterologous prime-boost (AZD1222-mRNA-1273 (A/M, light blue), AZD1222-BNT162b2 (A/P, light green) or vector-based (AZD1222-AZD1222 (A/A, green), Ad26.CoV2.S (J, grey))vaccination schemes in the mix and match cohort. SARS-CoV-2 RBD WT (Figure 3) is again shown for clarity and comparison. Data is shown as box and whisker plots. Boxes represent medians, 25th and 75th percentiles and whiskers show the largest and smallest non-outlier values based on 1.5 IQR calculation. Outlier values are shown in clear circles. The threshold for non-responsive samples (ACE2 binding inhibition less than 20%) is shown as dotted line. All samples below this can be considered non-responsive. Number of samples per vaccination scheme are stated below the figure.

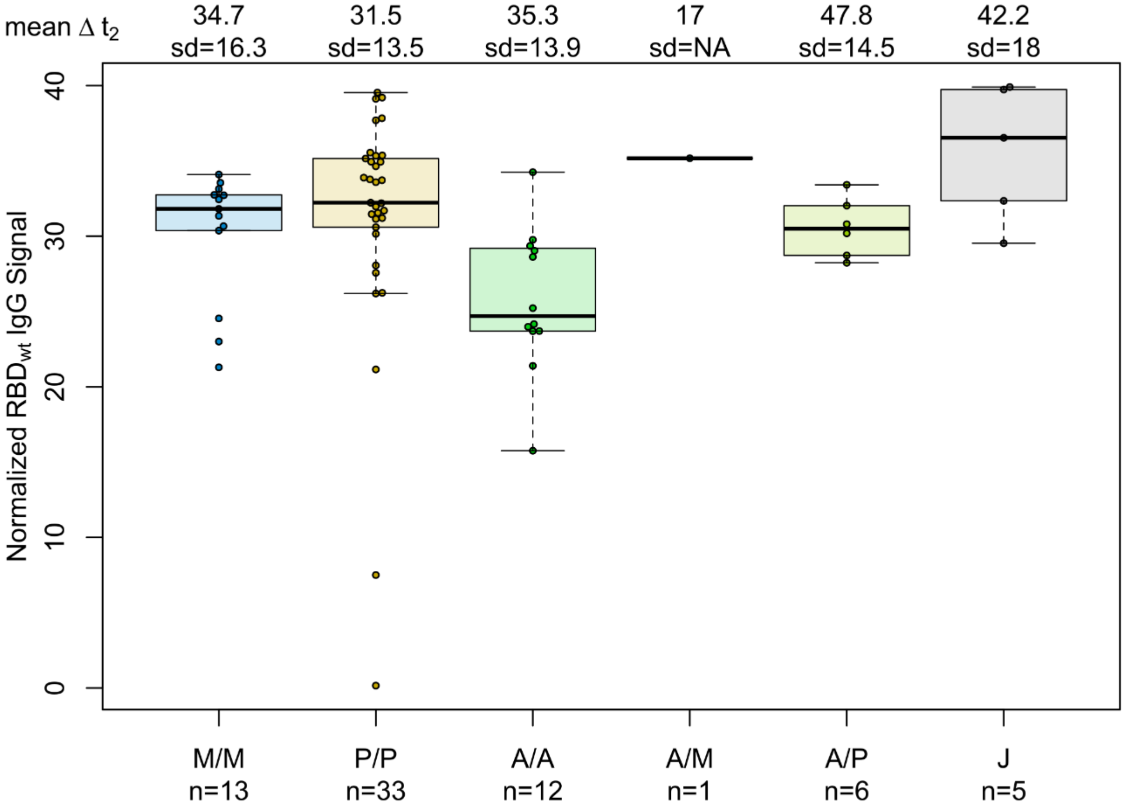


**Fig. S3. SARS-CoV-2 vaccination boosts humoral response among recovered individuals independent of vaccination scheme.**

Differences in vaccination responses of recovered previously SARS-CoV-2 infected individuals from our mix and match cohort were analysed using MULTICOV-AB. SARS-CoV-2 infection status was based on a previous self-reported positive PCR/antigen test or a MULTICOV-AB nucleocapsid IgG normalisation ratio above 1. Samples were split according to vaccination scheme (homologous mRNA (mRNA-1273 (M/M, blue), BNT162b2 (P/P, orange)), heterologous prime-boost (AZD1222-mRNA-1273 (A/M, light blue), AZD1222-BNT162b2 (A/P, light green) or vector-based (AZD1222-AZD1222 (A/A, green), Ad26CoV2.S (J, grey)). Boxes represent medians, 25th and 75th percentiles and whiskers show the largest and smallest non-outlier values based on 1.5 IQR calculation. When less than 5 were available per group, only the median is indicated by a line. Time between sampling and full vaccination is displayed as mean and SD for each group. Number of samples per vaccination scheme are stated below the figure. Results of a formal statistical comparison of recovered-vaccinated and SARS-CoV-2 naïve-vaccinated individuals is shown in Table S6.

**Table S1.** Comorbidities in study participants (n. a.: not applicable; NA: not available; CVD: cardiovascular disease).

| Sample cohort (n) | SARS-CoV-2 infection status (n) * | ΔT post-vaccination (days) | Vaccine (n) | Hyper-tension (n, %) | CVD (n, %) | Diabetes (n, %) | Lung disease (n, %) | Cancer (n, %) | Immuno-suppression (n, %) |
|---|---|---|---|---|---|---|---|---|---|
| **Mix and match (1470)** | + (70) | 7-65 | M/M (13) | 4 (30.8) | 1 (7.69) | 2 (15.4) | 0 (0) | 0 (0) | 0 (0) |
| | | | P/P (33) | 12 (36.0) | 4 (12.0) | 3 (9.10) | 0 (0) | 0 (0) | 1 (3.0) |
| | | | A/A (33) | 7 (58.3) | 3 (25.0) | 2 (16.67) | 1 (8.30) | 1 (8.30) | 2 (16.7) |
| | | | A/M (1) | 1 (100) | 0 (0) | 0 (0) | 0 (0) | 0 (0) | 0 (0) |
| | | | A/P (6) | 3 (50.0) | 1 (16.7) | 0 (0) | 1 (16.7) | 0 (0) | 0 (0) |
| | | | J (5) | 2 (40.0) | 0 (0) | 0 (0) | 0 (0) | 0 (0) | 1 (20.0) |
| | - (1400) | | M/M (272) | 84 (30.9) | 23 (8.46) | 13 (4.78) | 17 (6.25) | 2 (0.74) | 11 (4.04) |
| | | | P/P (734, 4 NA) | 263 (35.8) | 101 (13.8) | 64 (8.72) | 59 (8.04) | 20 (2.72) | 37 (5.04) |
| | | | A/A (228) | 91 (39.9) | 25 (11.1) | 25 (11.0) | 12 (5.26) | 2 (0.88) | 9 (3.95) |
| | | | A/M (24) | 12 (50.0) | 4 (16.7) | 2 (8.34) | 0 (0) | 1 (4.16) | 0 (0) |
| | | | A/P (114) | 43 (37.7) | 16 (14.0) | 10 (8.77) | 10 (8.77) | 4 (3.51) | 6 (5.26) |
| | | | J (24) | 9 (37.50) | 2 (8.30) | 2 (8.30) | 2 (8.30) | 1 (4.16) | 2 (8.30) |
| **Time points (597)** | - | 5-12 | P/P (107) | 50 (46.7) | 16 (15.0) | 6 (5.61) | 3 (2.80) | 0 (0) | 7 (6.50) |
| | | | M/M (40) | 13 (32.50) | 3 (7.50) | 3 (7.50) | 1 (2.50) | 0 (0) | 1 (2.50) |
| | | 26-30 | P/P (102; 1 NA) | 31 (30.4) | 15 (14.7) | 11 (10.8) | 12 (11.8) | 0 (0) | 0 (0) |
| | | | M/M (8) | 3 (37.5) | 1 (12.5) | 0 (0) | 0 (0) | 0 (0) | 0 (0) |
| | | 54-58 | P/P (92) | 37 (40.2) | 14 (15.2) | 7 (7.61) | 0 (0) | 2 (2.17) | 4 (4.35) |
| | | | M/M (22) | 2 (9.09) | 0 (0) | 2 (9.09) | 2 (9.09) | 0 (0) | 1 (4.55) |
| | | 94-103 | P/P (139) | 59 (42.5) | 18 (13.0) | 12 (8.63) | 1 (0.72) | 7 (5.04) | 0 (0) |
| | | | M/M (7) | 3 (42.9) | 1 (14.3) | 1 (14.3) | 1 (14.3) | 1 (14.3) | 0 (0) |
| | | 129-146 | P/P (38) | 19 (50.0) | 9 (23.7) | 7 (18.4) | 4 (10.5) | 2 (5.26) | 4 (10.5) |
| | | | M/M (5) | 3 (60.0) | 2 (40.0) | 1 (20.0) | 1 (20.0) | 0 (0) | 0 (0) |
| | | 176-203 | P/P (36) | 5 (13.9) | 2 (5.56) | 1 (2.78) | 2 (5.56) | 0 (0) | 3 (8.34) |
| | | | M/M (0) | n. a. | n. a. | n. a. | n. a. | n. a. | n. a. |
| **Longi-tudinal (90)** | - | 1st sample | P/P (90) | 35 (39.8) | 15 (17.1) | 11 (12.5) | 4 (4.6) | 2 (2.22) | 3 (3.41) |
| | | 2nd sample | | 33 (36.7) | 12 (13.3) | 8 (8.89) | 3 (3.33) | 1 (1.11) | 9 (10.0) |

Different vaccines and combinations are abbreviated as follows: M/M (two-dose mRNA-1273), P/P (two-dose BNT162b2), A/A (two-dose AZD1222), A/M (first dose AZD1222, second dose mRNA-1273), A/P (first dose AZD1222, second dose BNT162b2) and J (one-dose Ad26.CoV2.S). The time points sample cohort contains only homologous BNT162b2 and mRNA-1273 samples. The longitudinal sample cohort contains only homologous BNT162b2 samples.

* based on self-reported positive PCR/antigen test result at study centre visit and/or MULTICOV-AB nucleocapsid IgG S/CO ratio above 1

**Table S2.** MULTICOV-AB antigen panel.

| Virus | Antigen | Manufacturer | Product number |
|---|---|---|---|
| SARS-CoV-2 | Spike Trimer | NMI | - |
| SARS-CoV-2 | RBD B.1 (wild-type) | NMI | - |
| SARS-CoV-2 | Nucleocapsid | Aalto | 6404-b |
| SARS-CoV-2 | RBD B.1.1.7 (Alpha) | NMI | |
| SARS-CoV-2 | RBD B.1.351 (Beta) | NMI | |
| SARS-CoV-2 | RBD P.3 (Gamma) | NMI | |
| SARS-CoV-2 | RBD B.1.617.2 (Delta) | NMI | |
| hCoV-OC43 | S1 domain | NMI | - |
| hCoV-OC43 | Nucleocapsid | NMI | - |
| hCoV-HKU1 | S1 domain | NMI | - |
| hCoV-HKU1 | Nucleocapsid | NMI | - |
| hCoV-NL63 | S1 domain | NMI | - |
| hCoV-NL63 | Nucleocapsid | NMI | - |
| hCoV-229E | S1 domain | NMI | - |
| hCoV-229E | Nucleocapsid | NMI | - |

List of antigens included in MULTICOV-AB as well as their manufacturer and, if applicable, their product number.

**Table S3.** ACE2-RBD competition antigen panel.

| **Virus** | **Antigen** | **Manufacturer** | **Amino acid exchanges in RBD** |
|---|---|---|---|
| SARS-CoV-2 | RBD B.1 (wild-type) | NMI | - |
| SARS-CoV-2 | RBD B.1.1.7 (Alpha) | NMI | N501Y |
| SARS-CoV-2 | RBD B.1.351 (Beta) | NMI | N501Y, E484K, K417N |
| SARS-CoV-2 | RBD P.1 (Gamma) | NMI | N501Y, E484K, K417T |
| SARS-CoV-2 | RBD B.1.617.2 (Delta) | NMI | T478K, L452R |

List of antigens included in ACE2-RBD competition assay as well as their manufacturer, and the mutations covered within the RBD.

**Table S4.** Impact of confounders on ACE2 inhibition response towards SARS-CoV-2 wild-type and VoC RBDs.

| **A. WT ACE2 binding inhibition**, logit scale | **P/P, n=734** | | **M/M, n=272** | | **A/P, n=114** | | **A/A, n=228** | |
|---|---|---|---|---|---|---|---|---|
| | Est (sd) | p-value | Est (sd) | p-value | Est (sd) | p-value | Est (sd) | p-value |
| Intercept | 3.50 (0.231) | --- | 3.39 (0.330) | --- | 3.07 (0.538) | --- | -0.52 (0.496) | --- |
| Male | -0.25 (0.103) | 0.014 | -0.05 (0.149) | 0.752 | -0.33(0.267) | 0.218 | 0.30(0.152) | 0.050 |
| Age (per 1 year) | -0.03 (0.003) | <0.001 | 0.00 (0.005) | 0.431 | -0.01 (0.010) | 0.251 | 0.00(0.008) | 0.960 |
| Days post-vaccination (ΔT**)** | | | | | | | | |
| 7-27 | Ref. | | Ref. | | Ref. | | Ref. | |
| 28-65 | -0.69 (0.104) | <0.001 | -0.97 (0.169) | <0.001 | -0.74 (0.260) | 0.005 | -0.10(0.188) | 0.584 |
| Comorbidities | | | | | | | | |
| Cardiovascular | -0.12(0.158) | 0.454 | -0.54 (0.309) | 0.083 | -0.03(0.424) | 0.937 | 0.02 (0.241) | 0.943 |
| Hypertension | 0.19 (0.119) | 0.109 | -0.13 (0.180) | 0.480 | -0.08(0.301) | 0.789 | 0.11(0.161) | 0.485 |
| Diabetes | -0.04 (0.185) | 0.808 | 0.58(0.386) | 0.136 | 0.84(0.501) | 0.095 | -0.15(0.242) | 0.547 |
| Lung disease* | -0.03(0.187) | 0.853 | 0.53(0.324) | 0.104 | -0.87(0.513) | 0.091 | -0.11(0.349) | 0.758 |
| Cancer or Immunosuppression* | -0.26(0.192) | 0.176 | 0.29(0.362) | 0.418 | 0.22(0.508) | 0.662 | -0.61(0.368) | 0.099 |

| **B. Alpha ACE2 binding inhibition**, logit scale | **P/P, n=734** | | **M/M, n=272** | | **A/P, n=114** | | **A/A, n=228** | |
|---|---|---|---|---|---|---|---|---|
| | Est (sd) | p-value | Est (sd) | p-value | Est (sd) | p-value | Est (sd) | p-value |
| Intercept | 2.50(0.211) | --- | 2.64(0.301) | --- | 2.26(0.465) | --- | -0.77(0.420) | --- |
| Male | -0.22(0.083) | 0.008 | -0.02(0.130) | 0.862 | -0.27(0.226) | 0.228 | 0.24(0.126) | 0.060 |
| Age (per 1 year) | -0.02(0.003) | <0.001 | -0.01(0.004) | 0.218 | -0.01(0.009) | 0.270 | 0.00(0.006) | 0.826 |
| Days post-vaccination (ΔT) | | | | | | | | |
| 7-27 | Ref. | | Ref. | | Ref. | | Ref. | |
| 28-65 | -0.56(0.083) | <0.001 | -0.89(0.147) | <0.001 | -0.58(0.219) | 0.008 | -.03(0.155) | 0.842 |
| Comorbidities | | | | | | | | |
| Cardiovascular | -0.10(0.125) | 0.434 | -0.42(0.266) | 0.112 | -0.02(0.361) | 0.961 | 0.04(0.202) | 0.842 |
| Hypertension | 0.16(0.095) | 0.087 | -0.10(0.156) | 0.524 | -0.06(0.255) | 0.800 | 0.05(0.134) | 0.715 |
| Diabetes | -0.03(0.149) | 0.828 | 0.41(0.332) | 0.216 | 0.81(0.425) | 0.057 | -0.05(0.204) | 0.815 |
| Lung disease* | -0.03(0.149) | 0.851 | 0.40(0.281) | 0.156 | -0.78(0.433) | 0.073 | -0.08(0.288) | 0.789 |
| Cancer or Immunosuppression* | -0.15(0.153) | 0.343 | 0.21(0.314) | 0.510 | 0.15(0.428) | 0.724 | -0.45(0.302) | 0.139 |

| **C. Beta ACE2 binding inhibition**, logit scale | **P/P, n=734** | | **M/M, n=272** | | **A/P, n=114** | | **A/A, n=228** | |
|---|---|---|---|---|---|---|---|---|
| | Est (sd) | p-value | Est (sd) | p-value | Est (sd) | p-value | Est (sd) | p-value |
| Intercept | 1.27(0.259) | --- | 1.55(0.309) | --- | 1.12(0.420) | --- | -1.18(0.432) | --- |
| Male | -0.19(0.070) | 0.006 | -0.08(0.105) | 0.448 | -0.27(0.180) | 0.131 | 0.11(0.116) | 0.337 |
| Age (per 1 year) | -0.02(0.002) | <0.001 | 0.00(0.004) | 0.170 | -0.01(0.007) | 0.268 | 0.00(0.006) | 0.584 |
| Days post-vaccination (ΔT) | | | | | | | | |
| 7-27 | Ref. | | Ref. | | Ref. | | Ref. | |
| 28-65 | -0.44(0.070) | <0.001 | -0.86(0.119) | <0.001 | -0.59(0.175) | 0.001 | 0.07(0.144) | 0.645 |
| Comorbidities | | | | | | | | |
| Cardiovascular | 0.00(0.106) | 0.971 | -0.37(0.216) | 0.088 | -0.04(0.294) | 0.899 | 0.06(0.181) | 0.741 |
| Hypertension | 0.08(0.081) | 0.306 | -0.12(0.126) | 0.323 | -0.02(0.205) | 0.923 | 0.01(0.122) | 0.940 |
| Diabetes | 0.01(0.124) | 0.917 | 0.24(0.270) | 0.380 | 0.48(0.345) | 0.165 | 0.09(0.180) | 0.636 |
| Lung disease* | -0.08(0.126) | 0.520 | 0.39(0.227) | 0.087 | -0.54(0.346) | 0.117 | 0.10(0.268) | 0.705 |
| Cancer or Immunosuppression* | -0.19(0.130) | 0.155 | 0.13(0.254) | 0.606 | 0.27(0.340) | 0.426 | -0.40(0.286) | 0.163 |

| **D. Gamma ACE2 binding inhibition,** logit scale | **P/P, n=734** | | **M/M, n=272** | | **A/P, n=114** | | **A/A, n=228** | |
|---|---|---|---|---|---|---|---|---|
| | Est (sd) | p-value | Est (sd) | p-value | Est (sd) | p-value | Est (sd) | p-value |
| Intercept | 1.31(0.169) | --- | 1.64(0.250) | --- | 1.24(0.377) | --- | -1.05(0.313) | --- |
| Male | -0.20(0.067) | 0.003 | -0.06(0.109) | 0.612 | -0.36(0.183) | 0.049 | 0.13(0.092) | 0.165 |
| Age (per 1 year) | -0.02(0.002) | <0.001 | 0.00(0.004) | 0.222 | -0.01(0.007) | 0.183 | 0.00(0.005) | 0.382 |
| Days post-vaccination (ΔT) | | | | | | | | |
| 7-27 | Ref. | | Ref. | | Ref. | | Ref. | |
| 28-65 | -0.46(0.068) | <0.001 | -0.91(0.124) | <0.001 | -0.52(0.177) | 0.004 | -0.03(0.114) | 0.769 |
| Comorbidities | | | | | | | | |
| Cardiovascular | -0.06(0.101) | 0.556 | -0.41(0.225) | 0.069 | -0.05(0.299) | 0.862 | 0.13(0.145) | 0.352 |
| Hypertension | 0.14(0.078) | 0.063 | -0.08(0.132) | 0.548 | 0.04(0.208) | 0.862 | 0.07(0.097) | 0.496 |
| Diabetes | -0.01(0.118) | 0.960 | 0.21(0.281) | 0.463 | 0.53(0.351) | 0.134 | 0.03(0.145) | 0.821 |
| Lung disease* | -0.04(0.121) | 0.770 | 0.27(0.237) | 0.252 | -0.62(0.351) | 0.076 | -0.13(0.212) | 0.554 |
| Cancer or Immunosuppression* | -0.17(0.125) | 0.178 | 0.12(0.265) | 0.654 | 0.17(0.344) | 0.626 | -0.45(0.225) | 0.047 |

| **E. Delta ACE2 binding inhibition**, logit scale | **P/P, n=734** | | **M/M, n=272** | | **A/P, n=114** | | **A/A, n=228** | |
|---|---|---|---|---|---|---|---|---|
| | Est (sd) | p-value | Est (sd) | p-value | Est (sd) | p-value | Est (sd) | p-value |
| Intercept | 2.34(0.218) | --- | 2.53(0.312) | --- | 2.16(0.468) | --- | -1.10(0.430) | --- |
| Male | -0.21(0.085) | 0.013 | -0.03(0.135) | 0.830 | -0.34(0.226) | 0.132 | 0.20(0.128) | 0.128 |
| Age (per 1 year) | -0.02(0.003) | <0.001 | 0.00(0.004) | 0.349 | -0.01(0.009) | 0.203 | 0.00(0.007) | 0.698 |
| Days post-vaccination ($\Delta$T) | | | | | | | | |
| 7-27 | Ref. | | Ref. | | Ref. | | Ref. | |
| 28-65 | -0.55(0.086) | <0.001 | -0.99(0.152) | <0.001 | -0.70(0.220) | 0.001 | 0.01(0.159) | 0.938 |
| Comorbidities | | | | | | | | |
| Cardiovascular | -0.10(0.130) | 0.445 | -0.52(0.276) | 0.058 | 0.12(0.365) | 0.744 | 0.09(0.203) | 0.641 |
| Hypertension | 0.15(0.099) | 0.125 | -0.11(0.162) | 0.501 | 0.01(0.256) | 0.957 | -0.04(0.136) | 0.762 |
| Diabetes | -0.01(0.152) | 0.969 | 0.42(0.345) | 0.227 | 0.68(0.429) | 0.116 | -0.01(0.204) | 0.953 |
| Lung disease* | -0.02(0.155) | 0.920 | 0.39(0.292) | 0.182 | -0.67(0.434) | 0.125 | -0.15(0.295) | 0.604 |
| Cancer or Immunosuppression* | -0.18(0.159) | 0.261 | 0.17(0.326) | 0.603 | 0.06(0.428) | 0.888 | -0.50(0.311) | 0.111 |

ACE2 binding inhibition towards SARS-CoV-2 wild-type (A) and Alpha (B), Beta (C), Gamma (D), Delta (E) VoC in the mix and match sample cohort was modelled to assess the role of confounders (see methods for model details). Four samples (P/P) had missing information on comorbidities and were excluded from the analysis. Est=estimate, sd=standard deviation. Categories denoted with * were merged in the sensitivity analysis including interaction terms with time post-vaccination due to limited sample sizes (selected results reported in the main text).

.

**Table S5.** Statistical comparison of antibody titres between vaccination schemes.

| Comparison X | vs. Y | Full-length Spike trimer p'' (p-value) | RBD p'' (p-value) | S1 p'' (p-value) | S2 p'' (p-value) |
|---|---|---|---|---|---|
| J | M/M | 0.91 (< 0.001) * | 0.93 (<0.001) * | 0.96 (<0.001) * | 0.86 (< 0.001) * |
| | P/P | 0.89 (<0.001) * | 0.90 (<0.001) * | 0.92 (<0.001) * | 0.79 (< 0.001) * |
| | A/M | 0.89 (<0.001) * | 0.91 (<0.001) * | 0.97 (<0.001) * | 0.93 (<0.001) * |
| | A/P | 0.90 (<0.001) * | 0.92 (<0.001) * | 0.95 (<0.001) * | 0.88 (<0.001) * |
| | A/A | 0.63 (<0.080) | 0.74 (<0.001) * | 0.86 (<0.001) * | 0.81 (<0.001) * |
| A/A | M/M | 0.96 (<0.001) * | 0.94 (<0.001) * | 0.93 (<0.001) * | 0.57 (0.013) * |
| | P/P | 0.91 (<0.001) * | 0.87 (<0.001) * | 0.83 (<0.001) * | 0.39 (<0.001) * |
| | A/M | 0.93 (<0.001) * | 0.91 (<0.001) * | 0.93 (<0.001) * | 0.84 (< 0.001) * |
| | A/P | 0.93 (<0.001) * | 0.89 (<0.001) * | 0.91 (<0.001) * | 0.66 (< 0.001) * |
| A/P | M/M | 0.60 (0.002) * | 0.64 (<0.001) * | 0.55 (0.114) | 0.38 (< 0.001) * |
| | P/P | 0.42 (0.012) | 0.49 (0.608) | 0.34 (<0.001) * | 0.21 (< 0.001) * |
| | A/M | 0.59 (0.204) | 0.59 (0.155) | 0.69 (0.004) * | 0.77 (<0.001) * |
| A/M | M/M | 0.50 (0.982) | 0.53 (0.623) | 0.33 (0.014) * | 0.16 (<0.001) * |
| | P/P | 0.33 (0.014) | 0.40 (0.085) | 0.18 (<0.001)* | 0.09 (<0.001) * |
| P/P | M/M | 0.69 (<0.001) * | 0.64 (<0.001) * | 0.72 (<0.001) * | 0.71 (<0.001) * |

Estimated probability (p'') that a random individual under one vaccination scheme in the mix and match sample cohort, has a larger titre than a random individual under another vaccination scheme. Brunner-Munzel was used to determine statistical significance with Holm's adjustment for multiple testing. For a comparison of a scheme X with a scheme Y, a significant p'' larger than ½ means that the vaccination-induced titres under Y tend to be higher and a significant p'' smaller than ½ means that the vaccination-induced titres under X tend to be higher.

**Table S6**. Statistical comparison of RBD antibody titres and wild-type ACE2 binding inhibition in recovered-vaccinated and SARS-CoV-2 naïve-vaccinated individuals for different vaccination schemes

| | **M/M**<br>**p" (p-value)** | **P/P**<br>**p" (p-value)** | **A/M**<br>**p" (p-value)** | **A/P**<br>**p" (p-value)** | **A/A**<br>**p" (p-value)** | **J**<br>**p" (p-value)** |
|---|---|---|---|---|---|---|
| RBD | 0.66<br>(0.074) | 0.79<br>(<0.001) | -- (only 1 convalescent) | 0.79<br>(<0.001) | 0.94<br>(<0.001) | 0.98<br>(<0.001) |
| AC2 binding inhibition against WT | 0.74<br>(0.016) | 0.85<br>(<0.001) | -- (only 1 convalescent) | 0.84<br>(0.004) | 0.97<br>(<0.001) | 0.98<br>(<0.001) |

Estimated probability (p'') that a random recovered-vaccinated individual has a larger titre than a random SARS-CoV-2 naïve vaccinated individual under the respective vaccination scheme. Statistical analysis was determined by Brunner-Munzel. A significant p'' larger than ½ means that the vaccination-induced titres tend to be higher in recovered-vaccinated individuals.
